# Inhibiting LRRK2 kinase activity protects against the pathology associated with the presenilin 1 Ile416Thr mutation in cholinergic neurons

**DOI:** 10.1101/2025.09.17.25336024

**Authors:** Nicolas Gomez-Sequeda, Marlene Jimenez-Del-Rio, Carlos Velez-Pardo

## Abstract

Familial Alzheimer’s disease (FAD) is an accelerated form of dementia that affects cholinergic neurons. Unfortunately, despite several efforts, no single drug or treatment has shown complete efficacy. Therefore, it is imperative to find effective therapeutic agents. Previously, it has been shown that the PSEN1 I416T variant induces FAD-like neuropathology in cholinergic-like neurons (ChLNs), which is characterized by intracellular accumulation of the A(beta) peptide, oxidation of the stress sensor protein DJ-1, abnormal phosphorylation of the TAU protein at residue S202/T205, loss of mitochondrial membrane potential (deltapsi(m)), and activation of the pro-apoptotic proteins TP53, c-JUN, PUMA, and cleaved caspase 3 (CC3). We report for the first time that PSEN1 I416T induces (i) abnormal phosphorylation of LRRK2 kinase at residue S935, concomitant with (ii) phosphorylated alpha-synuclein (aSyn) at residue S129 and abnormal accumulation of autophagosomes and atypical increase (alkaline) in vesicular lysosomal pH, thereby provoking a profound alteration in autophagy in ChLNs. Here, we also demonstrate for the first time that the potent LRRK2 inhibitor PF-06447475 (hereafter referred to as PF475) almost completely attenuated PSEN1 I416T-induced proteinopathy, oxidative stress (OS), and apoptosis and restored autophagy to an activity comparable to untreated wild-type (WT) ChLNs. Overall, PF475 restored the survival of mutant ChLNs. Since LRRK2 kinase inhibitors are promising therapeutic drugs for the treatment of Parkinson’s disease (PD), our findings suggest that LRRK2 may also be a potential target for therapeutic intervention in FAD.

## Introduction

Alzheimer’s disease (AD) is a form of dementia that affects memory, thinking, and behavior [1], triggered by specific structural and functional losses of cholinergic neuronal projections from the nucleus basalis of Meynert (Ch4) to the cortex and from the medial septal nucleus (Ch1) to the hippocampus [2–4]. AD is typically characterized by the deposition of amyloid plaques formed by the extracellular accumulation of Aβ42 (eAβ42), intracellular accumulation of the abnormally phosphorylated protein TAU, and neuronal and synaptic loss [5]. AD can be classified as familial AD (FAD, with an onset age of <64 years) or sporadic AD (SAD, with an onset age of >65 years). Interestingly, FAD is more severe, both clinically and neuropathologically, than SAD [6]. To date, at least three highly penetrant mutated genes, named amyloid precursor protein (*APP*, >100 variants), presenilin (*PSEN*) 1 (*PSEN1*, 350 variants), and *PSEN2* (80 variants) (https://www.alzforum.org/alterations), as well as polygenic risk variants, have been identified that are genetically associated with FAD [7,8]. Specifically, a missense mutation in the *PSEN1* gene, known as PSEN1 I416T, resulting from an isoleucine to threonine residue substitution at codon 416, has been identified to cause familial EOAD in large family communities in Colombia [9, 10]. Unfortunately, no effective therapeutic approaches are currently available for the delay or prevention of this type of familial AD. Therefore, alternative therapeutic strategies are needed. However, the development of such therapeutics has been hampered by the lack of a valid in vitro or in vivo model of FAD. In addition, the identification of target proteins associated with PSEN1 pathology has been a challenging task.

Our laboratory has shown that PSEN1 I416T ChLNs exhibit not only the typical high eAβ42/Aβ40 ratio and abnormal phosphorylation of TAU at pathogenic residues S202/T205, but also accumulation of intracellular Aβ (iAβ) fragments, oxidative stress (OS) as evidenced by oxidation of the sensor protein DJ-1 (i.e., DJ-1C106-SO_3_), phosphorylation of the transcription factor c-JUN at pathogenic residues S63/S73, loss of mitochondrial membrane potential (ΔΨm), expression of apoptosis -a type of regulated cell death markers such as TP53, cleaved caspase 3 (CC3), and irresponsiveness to acetylcholine (ACh)-induced Ca^2+^ ion influx as evidence of neuronal dysfunction [11]. Interestingly, several molecules of natural (e.g., curcumin) and synthetic chemical origin (e.g., SP600125, methylene blue, 4-phenylbutyric acid, tramiprosate) alone or in combination have almost completely reversed the neuropathologic features of PSEN1 I416T mutant ChLNs [12, 13]. All of these reagents work either as anti-amyloidogenic, anti-TAU, antioxidant, and/or specific target blockers. With the exception of JNK kinase [12], no other major regulatory protein has been identified that is associated with Aβ- and TAU-induced OS and apoptosis in the ChLNs model of FAD.

In addition to the aforementioned proteins, leucine-rich region kinase 2 (LRRK2), a multifunctional Parkinson’s disease (PD) kinase [14], was recently shown to be endogenously phosphorylated at residue S935 in PSEN1 E280A ChLNs [15]. In addition, phosphorylated alpha synuclein (p-αSyn) at the pathogenic residue S129 (pS129-αSyn) was also found concomitantly with pS935-LRRK2 in the same mutant ChLNs [15]. Because pS935-LRRK2 not only phosphorylates pS129-αSyn [16] but may also be involved in OS [17, 18], apoptosis [19, 20], and autophagy [21], among other cellular functions [22,23] in typical PD dopaminergic neurons, we suspected that the neuropathological hallmark of PD αSyn may coexist with AD and that LRRK2 protein may function in both diseases. However, the mechanism by which LRRK2 induces the coexistence of αSyn, Aβ, and TAU in mutant PSEN1 ChLNs is unknown. Therefore, elucidating the role of LRRK2 may be an important consideration in understanding the pathogenesis of FAD and refining future clinical trials. Given that inhibition of LRRK2 kinase protects nerve-like differentiated cells from OS-induced cell death [24–26], we theorized that specific inhibition of LRRK2 might protect mutant PSEN1 ChLNs from Aβ-induced OS and apoptosis.

To test this hypothesis, we obtained wild-type (WT) and PSEN1 I416T ChLNs derived from menstrual stromal cells (MenSCs), a valid model of FAD [27]. After treatment with the specific LRRK2 kinase inhibitor PF-06447475 (thereafter referred to as PF475 [28]), cellular and biochemical analysis showed that this inhibitor effectively (i) inhibited LRRK2 kinase and that it almost completely reestablished the (ii) proteinopathy (e.g., iAβ, p-TAU, αSyn), (iii) OS (e.g., DJ-1 C106-SO_3_; DFC-positive cells), (iv) apoptosis (e.g., loss of ΔΨm, TP53, PUMA, CC3), and (v) dysfunctional autophagy-lysosomal pathway (ALP, e.g., accumulation of autophagosomes, acidic pH lysosomal vacuolar activity) in PSEN1 I416T ChLNs. Taken together, our data suggest that the use of the PF475 agent might be beneficial for reducing iAβ-, TAU-, and αSyn-induced ChLNs damage in FAD.

## Methods

### Obtention of Menstrual Stromal Cells-derived Cholinergic-like neurons

The procedure for differentiation followed the guidelines described in ref. [29]. Menstrual blood was collected from two donors: a healthy individual (Tissue Bank Code, TBC #59962) and an asymptomatic patient with familial Alzheimer’s disease (FAD) (TBC #45000). The procedure of obtaining MenSCs occurred from multiple (2-3) menstrual cycle from March to July, 2021. The cells derived from the healthy donor were identified and characterized as Wild Type (WT) PSEN1, while the cells from the patient with FAD were identified and characterized as PSEN1 I416T, as reported previously [11]. Both donors signed writing informed consent forms, which were approved by the ethics committee of the Sede de Investigación Universitaria (SIU) at the University of Antioquia, Medellín, Colombia (Act 20-10-846). Consent was obtained from all participants in the research. The mesenchymal stem cells (MSCs) were initially seeded at a density of 1–1.5 × 10^4^ cells/cm² on plates coated with laminin using a standard culture medium (RCm) that included low-glucose DMEM and 10% fetal bovine serum (FBS) for 24 hours. After this period, the culture medium was switched to a cholinergic differentiation medium, known as Ch-N-Rm, which comprised DMEM/F-12 Nutrient Mixture (Gibco cat# 10565018, Grand Island, NY, USA), 1% fetal bovine serum (BSA), and a specific mix of nerve growth factors as cited elsewhere [30]. This process was maintained at 37 °C for 7 days. The transdifferentiated cells (85-100% double cholinergic marker Choline Acetyltransferase (ChAT)/ Vesicular Acetylcholine transporter (VAChT)-positive cells) were designated as WT PSEN1 or I416T ChLNs. After the initial transdifferentiation phase, the ChLN cells were cultured for an additional four days in regular culture medium (RCm) to eliminate any elements (e.g., growth factors) in Ch-N-Rm that might interfere with the experiment’s accuracy and data interpretation.

### Assay Protocol

PF-06447475 (PF475, Sigma-Aldrich cat# PZ0248), a selective LRRK2 inhibitor, was used at an optimal concentration of 1 µM. At this concentration, PF475 effectively inhibited the phosphorylation of LRRK2 kinase at serine 935 (S935), completely prevented the generation of reactive oxygen species (ROS), and significantly reversed all apoptosis signaling and oxidative stress markers induced by rotenone, restoring them to levels comparable to the control as shown by [15, 24]. For these analyses, the ChLN cells were divided into four groups: (i) untreated WT PSEN1; (ii) WT PSEN1 treated with PF475 (1 µM); (iii) untreated PSEN1 I416T; and (iv) PSEN1 I416T treated with PF475 (1 µM). The chemical inhibition assay was performed by pre-incubating the ChLN cells with PF475 (1 µM) for 4 days in regular culture medium (RCm).

### Immunofluorescence Analysis

The detection of markers associated with Alzheimer’s disease, oxidative stress, and cell death was conducted following the method outlined by [11], and theoretical considerations as described in Ref. [31]. Briefly, cells subjected to various treatments, as previously described, were initially fixed with 4% paraformaldehyde for 20 minutes, followed by permeabilization using 0.1% Triton X-100 and a blocking step with 10% bovine serum albumin (BSA). For staining, cells were incubated overnight at 4 °C with primary antibodies targeting several specific proteins, as detailed in Table 1. These antibodies include those for detecting the N-terminal amino acids of the Aβ peptide, total and phosphorylated tau for evaluating tau pathology, and oxidized DJ-1 for assessing oxidative stress. Antibodies against α-synuclein and LRRK2 were also used to evaluate protein aggregation and LC3-II to monitor autophagic flux. To analyze markers of apoptosis, cells were stained with primary antibodies conjugated to fluorescent dyes, such as those against PUMA, TP53, phospho-c-JUN, and a caspase-3 activity fluorescent probe. Additionally, to evaluate lysosomal activity, cells were incubated with LysoTracker Green® DND-26 in combination with LC3-II antibody staining to monitor autophagic vesicles. After thorough washing, the cells were incubated with appropriate secondary fluorescent antibodies, as specified in Table 1, to visualize primary antibody binding.

**Table 1.**
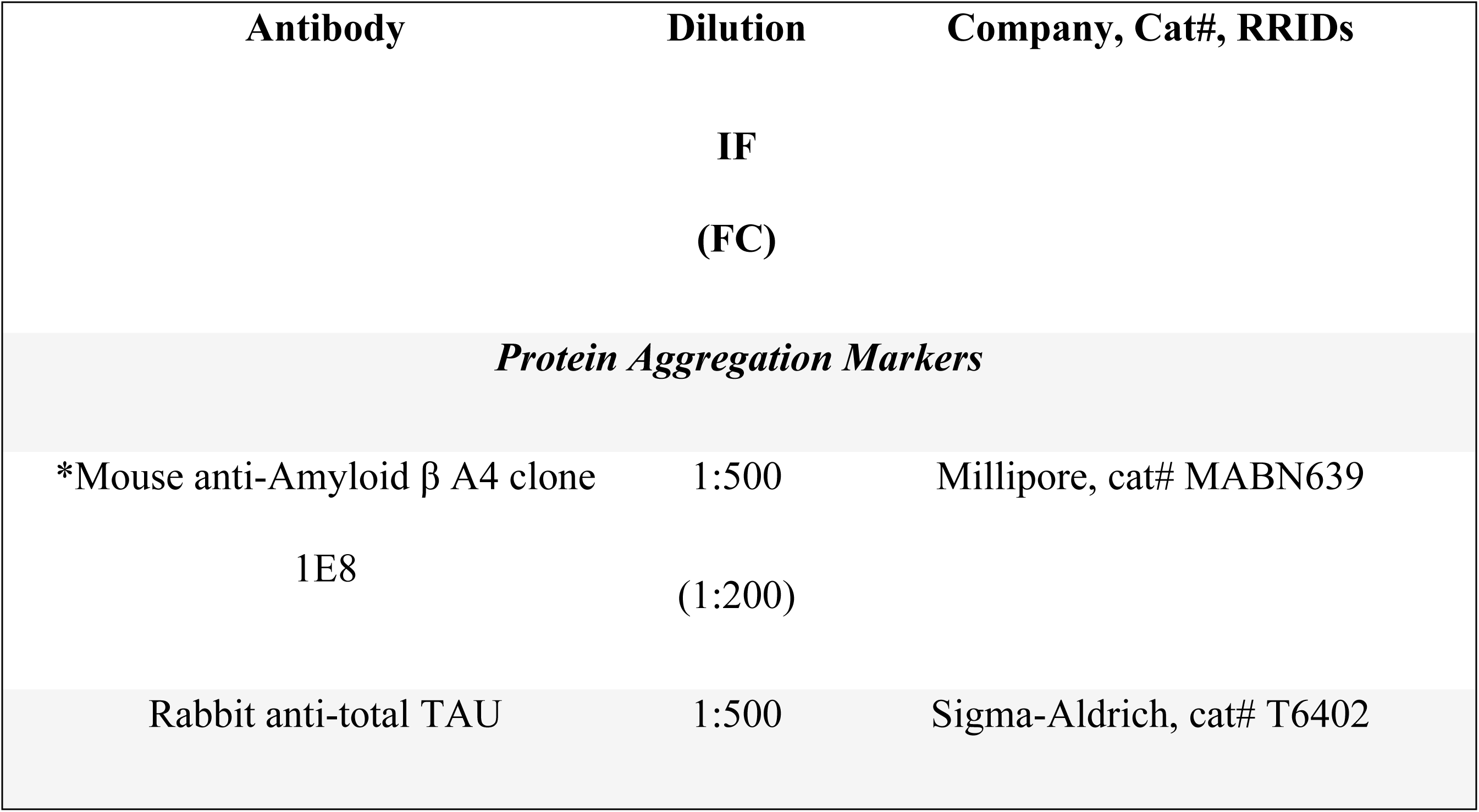
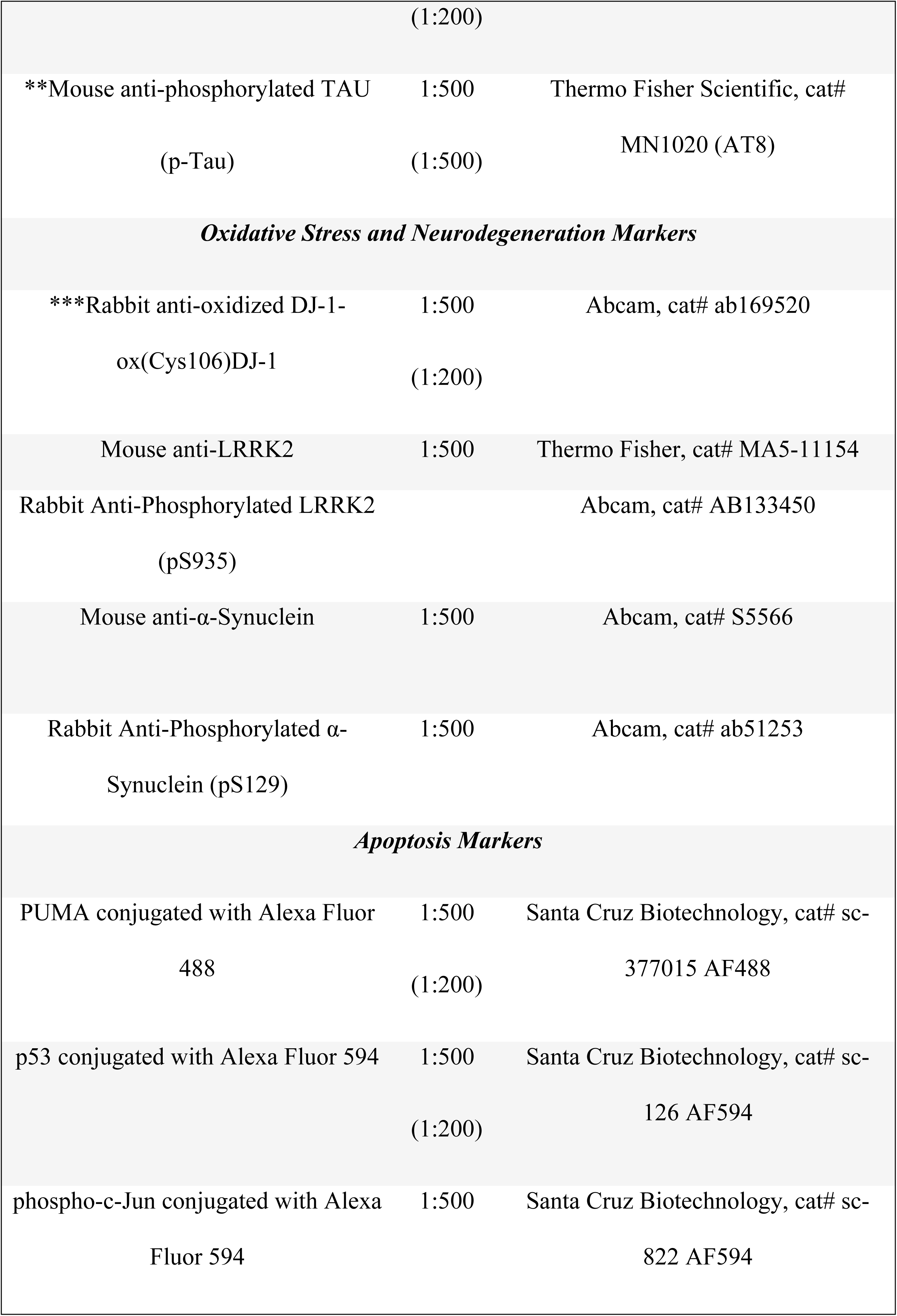
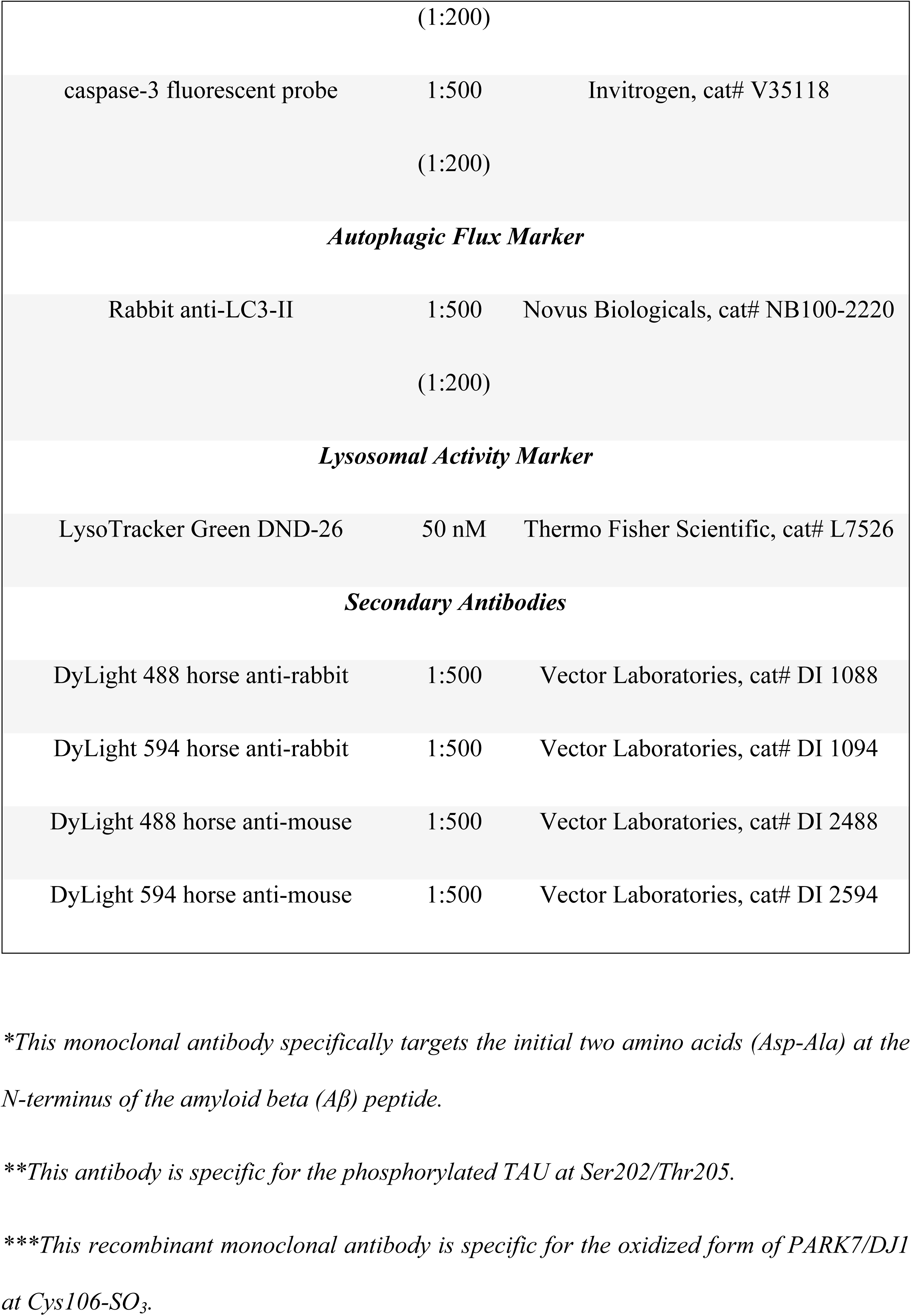
List of antibodies used for immunofluorescence (IF) staining and flow cytometry (FC) analysis of protein aggregation, oxidative stress, neurodegeneration, apoptosis, autophagy flux, and lysosomal activity.

### Flow Cytometry Analysis

After each treatment, cells were detached using trypsin and then centrifuged for 10 minutes at 2000 rpm. The cells were subsequently washed with phosphate-buffered saline (PBS) and fixed overnight in cold 96% ethanol. Following two additional washes with PBS, the cells were permeabilized for 30 minutes using a solution containing 0.2% Triton X-100 and 1.5% bovine serum albumin (BSA). The permeabilized cells were then incubated overnight at 4 °C with primary antibodies targeting various proteins, including markers for Alzheimer’s disease, oxidative stress, and apoptosis, as outlined in Table 1. After thorough rinsing to remove unbound primary antibodies, the cells were incubated with the appropriate secondary fluorescent antibodies, also detailed in Table 1, diluted to 1:500. Fluorescence was measured using a BD LSRFortessa II flow cytometer (BD Biosciences), with cells not treated with primary antibodies serving as the negative control. For each sample, data from 10,000 events were collected. The data was analyzed using FlowJo 10.8.1 software (TIBCO® Data Science) to generate quantitative figures and visualizations. Event analysis was performed by identifying cell populations that exceeded the baseline fluorescence of the negative control (measured at 488 nm or 594 nm), which facilitated the creation of density plots and histograms to visualize the distribution and intensity of fluorescence across different cell populations [32].

### Assessment of Intracellular Reactive Oxygen Species (ROS) by Fluorescence Microscopy and Flow Cytometry

To evaluate intracellular reactive oxygen species (ROS), we employed 2′,7′-dichlorofluorescein diacetate (5 μM, DCFH_2_-DA; Invitrogen). ChLNs were maintained in RCm for four days. Subsequently, cells (5 × 10³) were treated with DCFH2-DA for 30 minutes at 37 °C in the dark. Following this, cells were washed, and the fluorescence intensity of DCF was assessed using fluorescence microscopy. The nuclei were counterstained with Hoechst 33342 (0.5 μM). The experiment was conducted three times to ensure reliability. For a complementary analysis, cells (1 × 10⁴) were subjected to the same DCFH_2_-DA treatment and then analyzed using flow cytometry with an LSRFortessa (BD Biosciences). The quantitative data were analyzed with FlowJo 10.8.1 software. This assessment was also performed in triplicate, with both the experimenter and the flow cytometer analyst being blind to the experimental conditions.

### Evaluation of Mitochondrial Membrane Potential (ΔΨm) by Fluorescence Microscopy and Flow Cytometry

To assess mitochondrial membrane potential (ΔΨm), ChLNs were cultured in regular medium (RCm) for four days. Cells (5 × 10³) were then incubated with MitoTracker deep red dye (20 nM, final concentration; cat# M22426, Invitrogen, Waltham, MA, USA) for 20 minutes at room temperature in the dark. Following the incubation, cells were washed twice with PBS, and MitoTracker fluorescence intensity was analyzed using fluorescence microscopy. The nuclei were counterstained with Hoechst 33342 (0.5 μM). This procedure was repeated in three independent experiments to ensure consistency. For a complementary analysis, ChLNs (1 × 10⁴) were treated with the same MitoTracker dye and subsequently analyzed using an LSRFortessa flow cytometer (BD Biosciences). Data acquisition was performed for 10,000 events per sample, and the results were analyzed using FlowJo 10.8.1 software. The flow cytometry-based assessment was also repeated three times, with both the experimenter and the flow cytometer analyst being blind to the experimental conditions.

### Imaging and Quantitative Analysis of Fluorescence Microscopy Data

Optical and fluorescence microscopy images were acquired using a Zeiss Axio Vert.A1 microscope with an AxioCam Cm1 camera. For fluorescence microscopy, the Zeiss Axio Vert.A1 Fluorescence Microscope was employed. Fluorescence images were processed by converting them into 8-bit format and subtracting the background using Zen 3.4 blue edition software. Subsequent analysis was conducted with ImageJ software (http://imagej.nih.gov/ij/), utilizing a custom macro. Regions of interest (ROIs) were delineated around nuclei for analyzing transcription factors and apoptosis markers or around the entire cell body for cytoplasmic probes. Fluorescence intensity was measured by applying a uniform threshold across control and treatment conditions. The mean fluorescence intensity (MFI) was calculated by normalizing the total fluorescence to the number of nuclei, with a standard of 250 nuclei per analysis.

### Experimental Design and Statistical Analysis

Two vials of MSCs (WT PSEN1 and PSEN1 E280A) were thawed, cultured, and standardized to a density of 2.6 × 10⁴ cells/cm² before being pipetted into different wells of a 24-well plate. Cells, representing the biological and observational units [33], were randomly assigned to wells using a simple randomization method (sampling without replacement). Subsequently, wells, serving as the experimental units, were randomized to different treatments using the same method. Experiments were performed on three independent occasions (n = 3) blind to the experimenter and/or flow cytometer analyst [33]. The data from the three repetitions, i.e., independent experiments, were averaged, and a representative flow cytometry density or histogram plot from the three independent experiments was selected for illustrative purposes, whereas the bars in the quantification figures represent the mean ± SD and the three black dots show the data point of each experimental repetition. Based on the assumption that the experimental unit (i.e., the well) data comply with the independence of observations, the dependent variable is normally distributed in each treatment group (Shapiro-Wilk test), and variances are homogeneous (Levene’s test), the statistical significance was determined by one-way analysis of variance (ANOVA) followed by Tukey’s post hoc comparison calculated with GraphPad Prism 9 software. Differences between groups were only deemed significant when a p-value of <0.05 (*), <0.001 (**), and <0.001 (***). All data are illustrated as the mean ± S.D.

## Results

### Inhibitor LRRK2 PF475 inhibits the phosphorylation of LRRK2 kinase in PSEN1 I416T ChLNs

Because PSEN1 E280A ChLNs expressed endogenously phosphorylated pS935-LRRK2 concomitantly with pS129-αSyn, as a first approach, we evaluated whether LRRK2 kinase would be active in PSEN1 I416T and whether PF475 would block p-LRRK2. As shown in Figure 1, WT ChLNs expressed active pS935-LRRK2 at basal levels (Figure 1A), but the inhibitor reduced phosphorylated LRRK2 by −67% (Figure 1B). Interestingly, PSEN1 I416T ChLNs increased endogenously active phosphorylated LRRK2 by +144% (Fig. 1C) compared to untreated WT ChLNs (Fig. 1A). Upon exposure to PF475, pS935-LRRK2 in PSEN1 I416T was reduced by −54% (Fig. 1D) compared to untreated mutant ChLNs or reduced to basal levels (Fig. 1A vs. 1D). LRRK2 protein expression was nearly 100% in all neuronal cells (Figure 1A’-D’). Overall, PF475 reduced the ratio of pS935 LRRK2 (Fig. 1E) to total LRRK2 (Figures 1A’-D’, 1F) by approximately 3.2-fold and 2.2-fold in WT and mutant ChLNs, respectively (Fig. 1G). Similar results were obtained by fluorescence microscopy analysis (Figures 1H-N).

**Figure 1.**
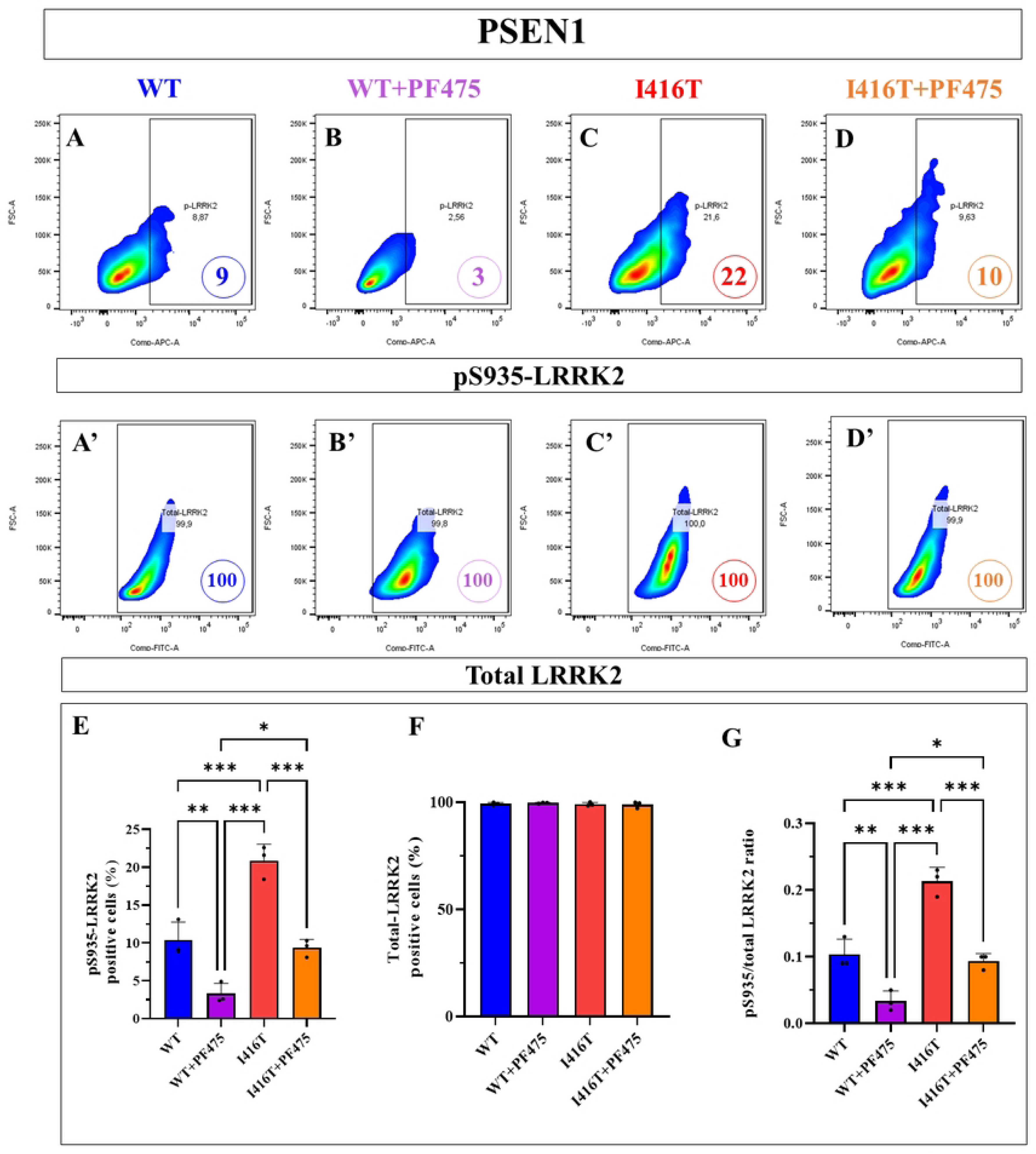
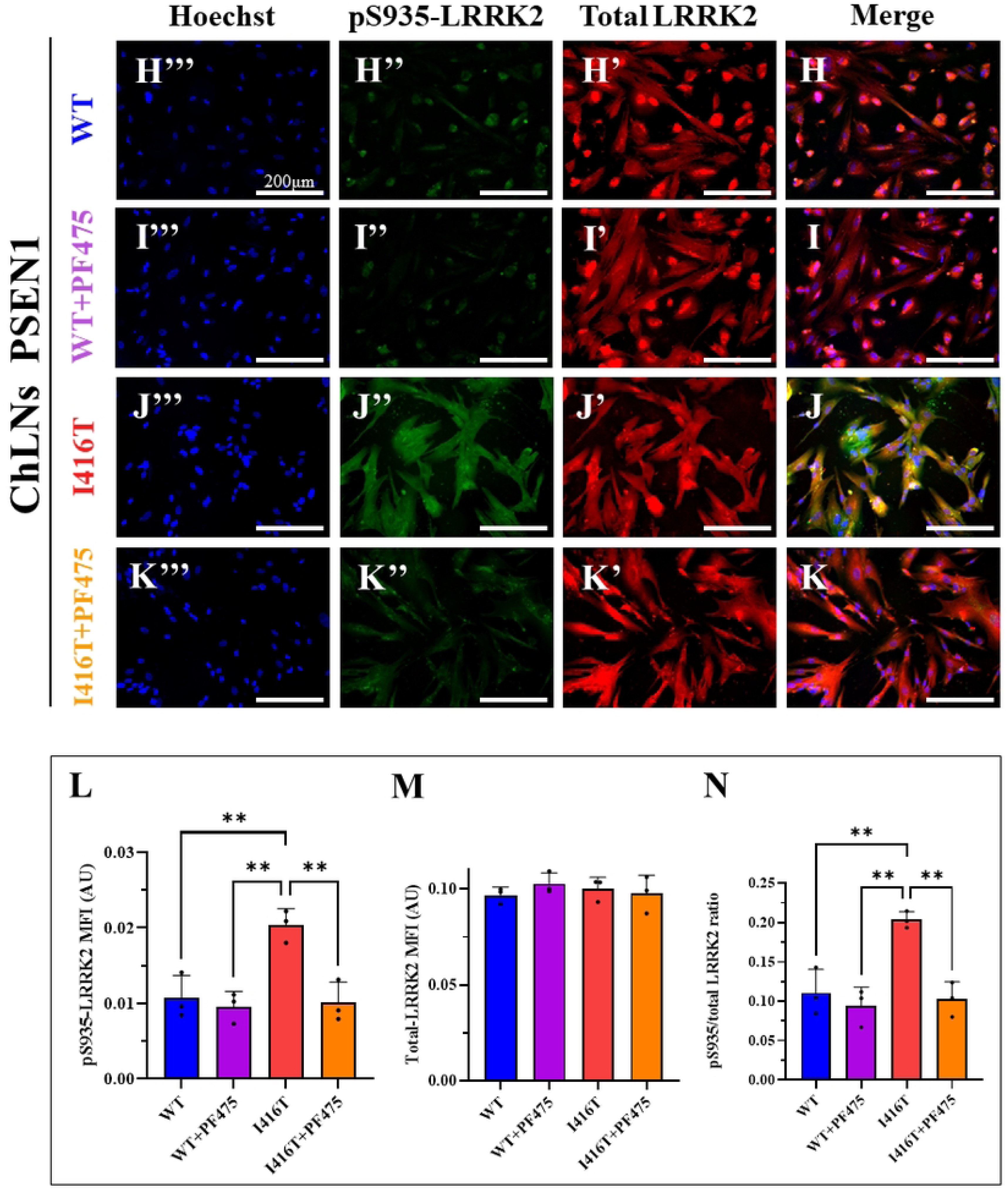
PF475 inhibits pS935-LRRK2 in wild-type (WT) PSEN1 and PSEN1 I416T Cholinergic-Like Neurons (ChLNs) reducing LRRK2 phosphorylation to basal levels. WT PSEN1 and I416T MenSCs were cultured in a cholinergic differentiation medium for 7 days, followed by 4 days in regular medium in the absence or presence of PF475 (1 μM). (A-D) Representative flow cytometry density plot with density contours analysis showing phosphorylated LRRK2-positive cells at residue S935 (A-D), total LRRK2-positive cells (A’-D’) in WT PSEN1 (A, B) or PSEN1 I416T ChLNs (C, D) in the absence (A, C) or presence (B, D) of PF475. (E-G) Quantification of pS935-LRRK2-positive cells (E), total LRRK2-positive cells (F), and the pS935/total LRRK2 ratio (G). (H-K) Representative immunofluorescence merge image showing pS935-LRRK2 in WT (H, I) or PSEN1 I416T ChLNs (J, K) in the absence (H, J) or presence (I, K) of PF475 stained with antibodies against total LRRK2 (H’-K’, red fluorescence), pS935-LRRK2 (H’’-K’’, green fluorescence). The nuclei were counterstained using Hoechst 33342 (H’’’-K’’’, blue fluorescence). (L-N) Mean fluorescence intensity (MFI) quantification of pS935-LRRK2 (L), total LRRK2 (M), and the pS935/total LRRK2 ratio (N). Figures represent one of three independent experiments (n = 3). Results are represented as mean ± standard deviation. Statistical significance was analyzed using one-way ANOVA followed by Tukey’s test *p < 0.05, **p < 0.01, ***p < 0.001. Images were taken at 200× magnification.

### PF475 inhibits the phosphorylation of αSynuclein (αSyn)

Since LRRK2 kinase directly or indirectly phosphorylates αSyn at the pathological residue S129, we investigated whether PSEN1 I416T expresses pS129-αSyn and whether PF475 blocks LRRK2-induced αSyn phosphorylation. Figure 2 shows that αSyn was phosphorylated at similar basal levels in untreated WT ChLNs (Figure 2A) or treated with PF475 (Figure 2B). While endogenous pS129-αSyn increased by +240% in PSEN1 I416T (Figure 2C) compared to untreated WT ChLNs, upon exposure to PF475, p-αSyn decreased by −59% in mutant neurons (Figure 2D) compared to untreated mutant ChLNs. Since total αSyn protein was nearly 100% in all neuronal cells (Figures 2A’-D’), PF475 reduced the ratio of pS129-αSyn (Figure 2E)/t-αSyn (Figure 2F) by 2.85-fold in PSEN1 I475T ChLNs (Figure 2G). Similar observations were obtained by fluorescence microscopy analysis (Figures 2H-N).

**Figure 2.**
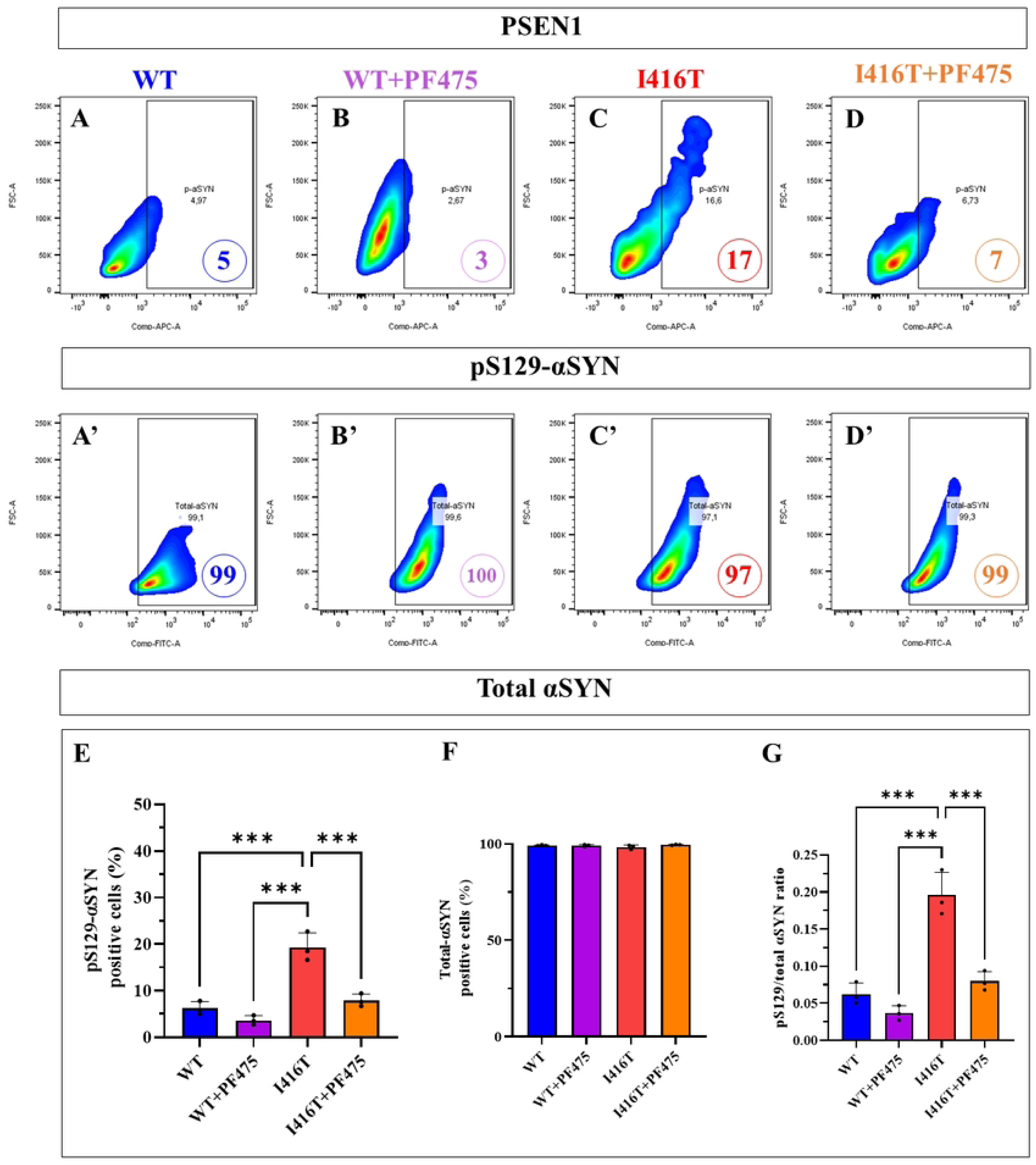
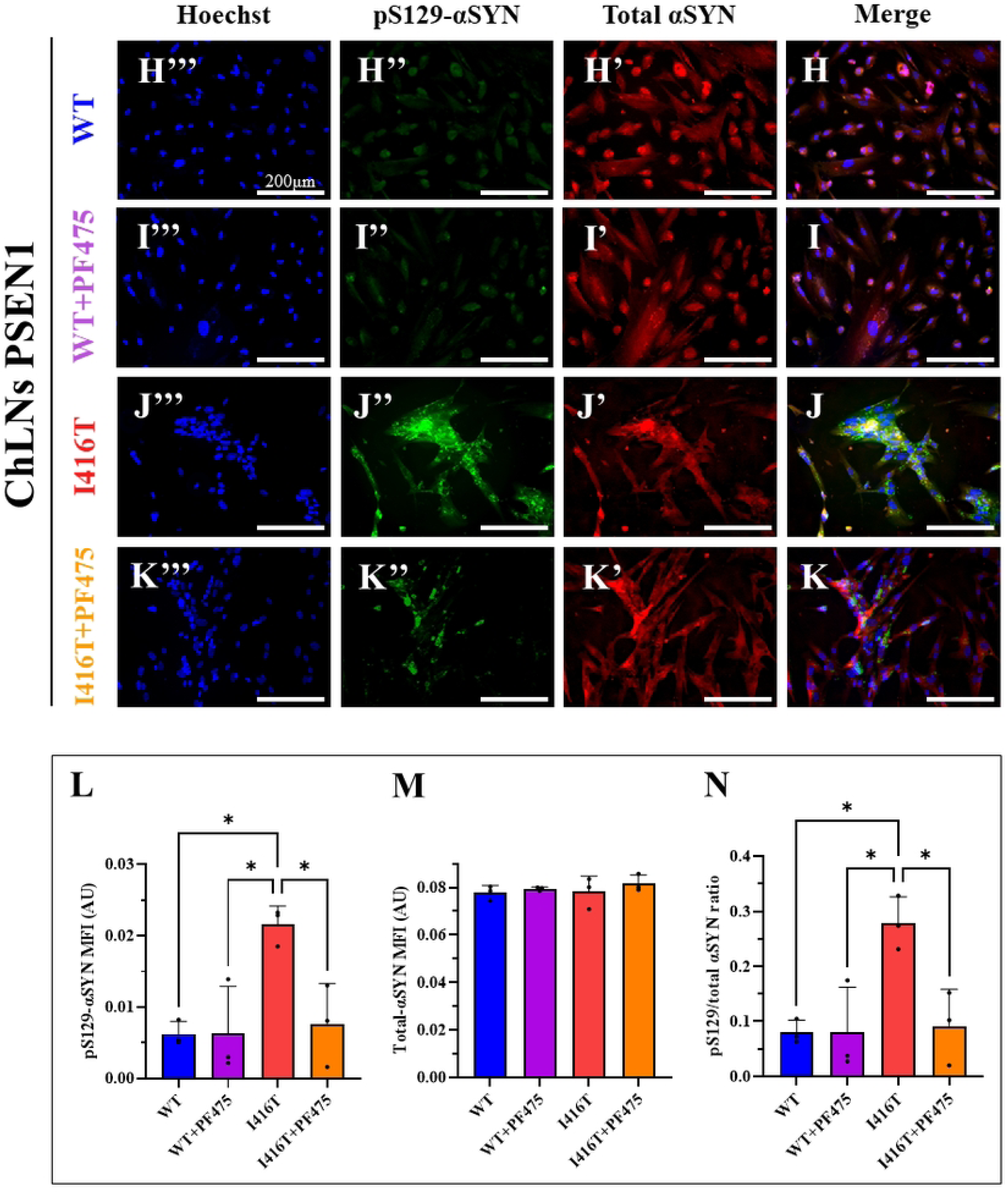
PF475 reduces pathological phosphorylation of αSynuclein (p-S129) in PSEN1 WT and I416T Cholinergic-Like Neurons (ChLNs). WT PSEN1 and I416T MenSCs were cultured in a cholinergic differentiation medium for 7 days, followed by 4 days in regular medium in the absence or presence of PF475 (1 μM). (A-D) Representative flow cytometry density plot with density contours analysis showing phosphorylated αSyn-positive cells at residue S129 (A-D), total αSyn-positive cells (A’-D’) in WT PSEN1 (A, B) or PSEN1 I416T ChLNs (C, D) in the absence (A, C) or presence (B, D) of PF475. (E-G) Quantification of pS129-αSyn-positive cells (E), total LRRK2-positive cells (F), and the pS129/total αSyn ratio (G). (H-K) Representative immunofluorescence merge image showing pS129-αSyn in WT (H, I) or PSEN1 I416T ChLNs (J, K) in the absence (H, J) or presence (I, K) of PF475 stained with antibodies against total αSyn (H’-K’, red fluorescence), pS129-αSyn (H’’-K’’, green fluorescence). The nuclei were counterstained using Hoechst 33342 (H’’’-K’’’, blue fluorescence). (L-N) Mean fluorescence intensity (MFI) quantification of pS129-αSyn (L), total αSyn (M), and the pS129/total αSyn ratio (N). Figures represent one of three independent experiments (n = 3). Results are represented as mean ± standard deviation. Statistical significance was analyzed using one-way ANOVA followed by Tukey’s test *p < 0.05, ***p < 0.001. Images were taken at 200× magnification.

### PF475 diminishes accumulation of iAβ, oxidization of protein DJ-1, and phosphorylated TAU protein in PSEN1 I416T ChLNs

We next sought to determine whether PF475 reduces the intracellular accumulation of Aβ (iAβ) and the oxidative stress marker oxDJ-1, which is a major target of ROS/H_2_O_2_ [34] and whether it blocks pS202/T205-TAU in mutant ChLNs [11]. As shown in Figure 3, while untreated WT ChLNs (Figures 3A and 3F) or inhibitor-treated ChLNs (Figure 3B and 3G) expressed similar basal levels of iAβ (Figure 3E) and oxDJ-1 (Figure 3J), mutant I416T ChLNs increased iAβ and oxDJ-1 by +560% (Figure 3C) and +660% (Figure 3H), respectively, compared to WT ChLNs (Figures 3E and 3J). Upon exposure to PF475, PSEN1 I416T dramatically reduced iAβ and oxDJ-1 by −73% (Figure 3D) and −78% (Figure 3I), respectively (Figures 3E and 3J). Similar observations were obtained by fluorescence microscopy analysis (Figures 3K-P).

**Figure 3.**
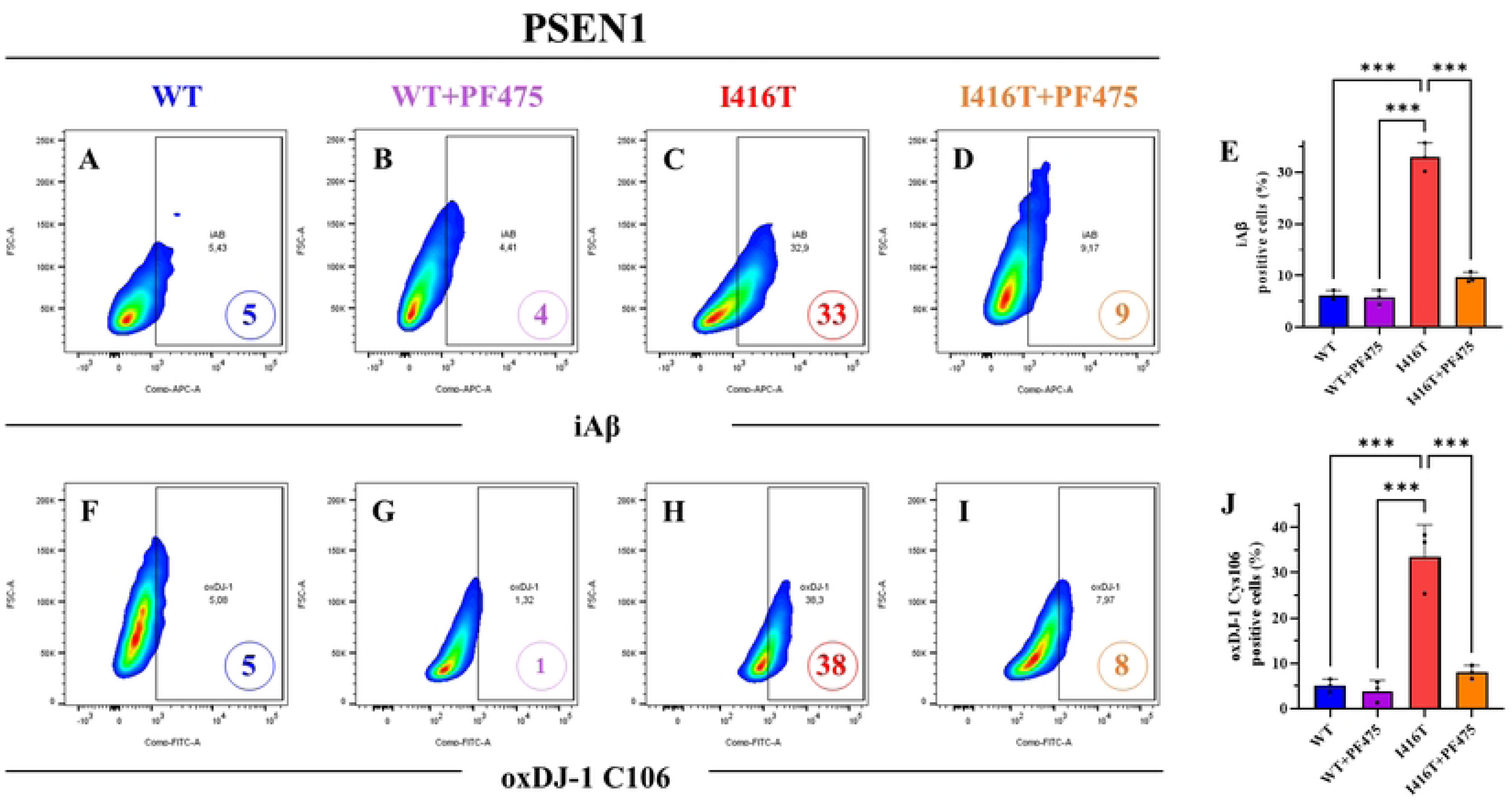
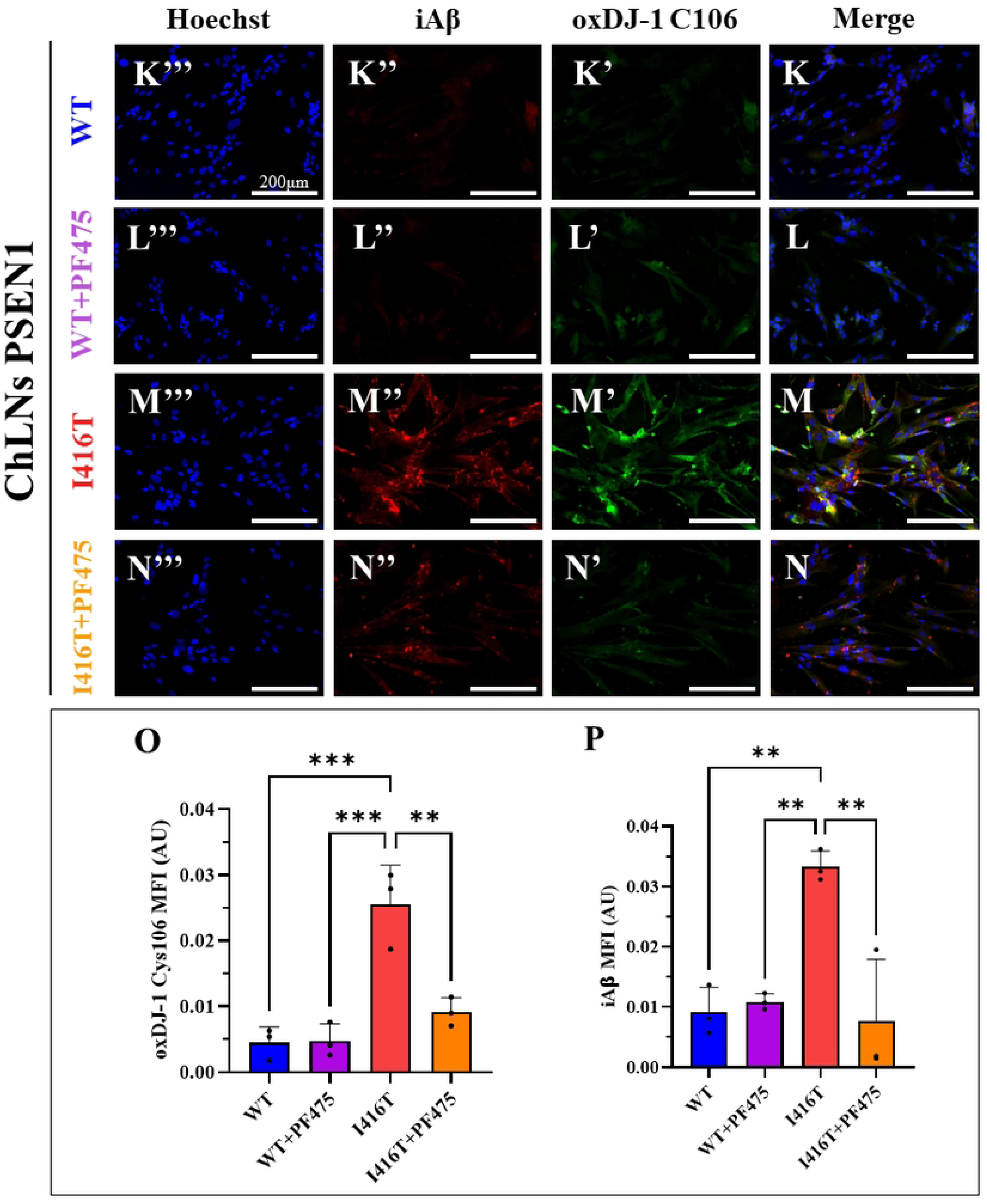
PF475 blocks ROS-induced oxidation of DJ-1 and intracellular Aβ accumulation in PSEN1 I416T Cholinergic-Like Neurons (ChLNs). WT PSEN1 and I416T MenSCs were cultured in a cholinergic differentiation medium for 7 days, followed by 4 days in regular medium in the absence or presence of PF475 (1 μM). (A-D) Representative flow cytometry density plot with density contours analysis showing iAβ- positive cells in WT PSEN1 (A, B) or PSEN1 I416T ChLNs (C, D) in the absence (A, C) or presence (B, D) of PF475. (E) Quantification of iAβ. (F-I) Representative flow cytometry density plot with density contours analysis showing oxDJ-1-positive cells in WT PSEN1 (F, G) or PSEN1 I416T ChLNs (H, I) in the absence (F, H) or presence (G, I) of PF475. (J) Quantification of oxDJ-1. (K-N) Representative immunofluorescence merge image showing oxDJ-1 and iAβ in WT (K, L) or PSEN1 I416T ChLNs (M, N) in the absence (K, M) or presence (L, N) of PF475 stained with antibodies against oxDJ-1 (K’-N’, green fluorescence), or iAβ (K’’-N’’, red fluorescence). The nuclei were counterstained using Hoechst 33342 (K’’’-N’’’, blue fluorescence). (O-P) Mean fluorescence intensity (MFI) quantification of oxDJ-1 (O) and iAβ (P). Figures represent one of three independent experiments (n = 3). Results are represented as mean ± standard deviation. Statistical significance was analyzed using one-way ANOVA followed by Tukey’s test **p < 0.01, ***p < 0.001. Images were taken at 200× magnification.

Like iAβ and oxDJ-1, PF475 decreased phosphorylated TAU (Figure 4). Indeed, p-TAU was expressed at similar basal levels in untreated (Figure 4A) or treated WT ChLNs (Figure 4B). PSEN1 I416T showed an increase of p-TAU by +580% (Fig. 4C), but upon PF475 treatment, p-TAU decreased by −240% (Figure 4D) in PSEN1 I416T compared to untreated mutant ChLNs (Figure 4E). Calculation of the proportion of phosphorylated TAU shows that PF475 reduced the ratio of p-TAU to total TAU protein (Figure 4A’-D’, 4F) by 2.75-fold in PSEN1 I416T (Figure 4G). Similar observations were obtained by fluorescence microscopy analysis (Figures 4H-N).

**Figure 4.**
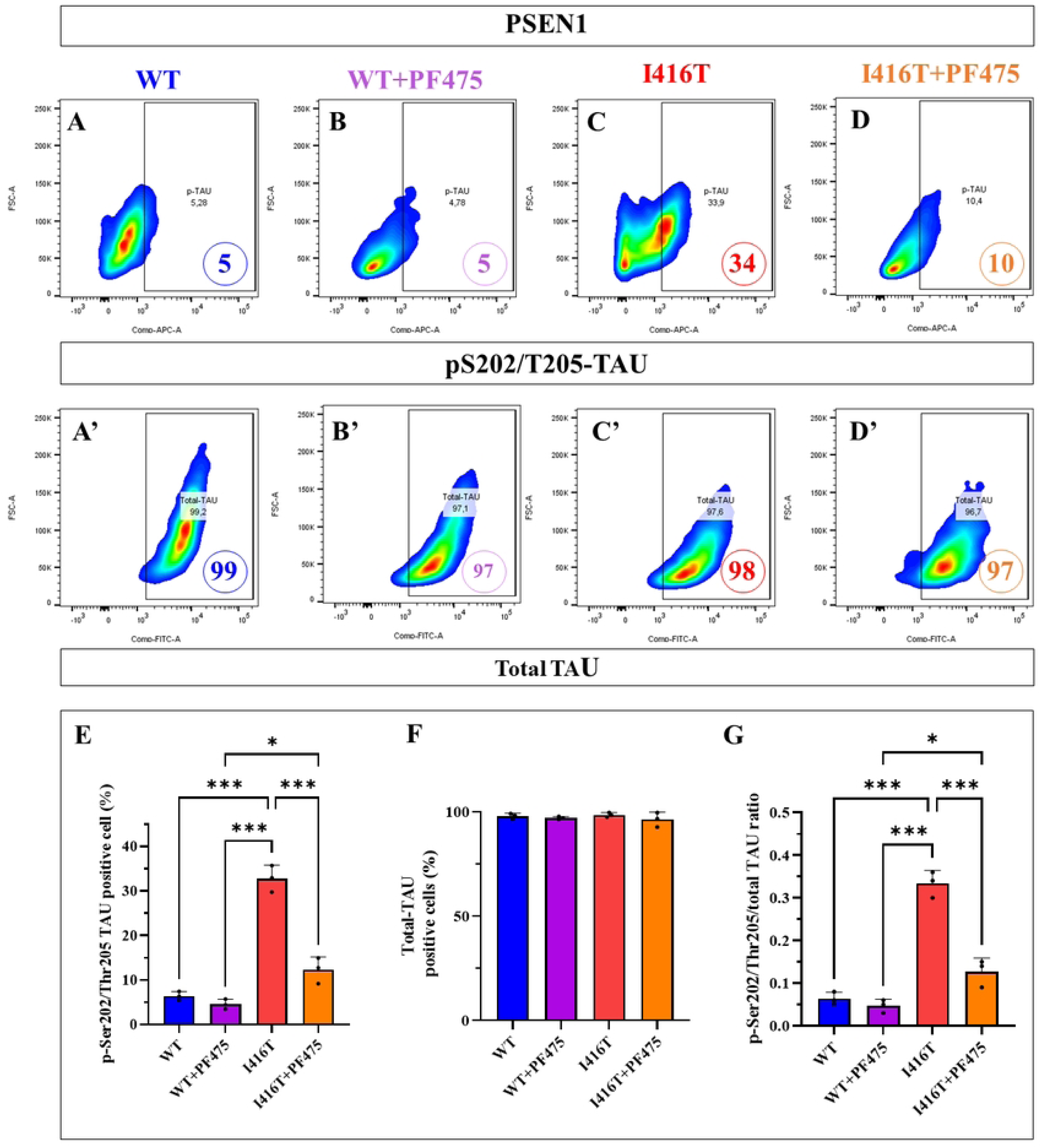
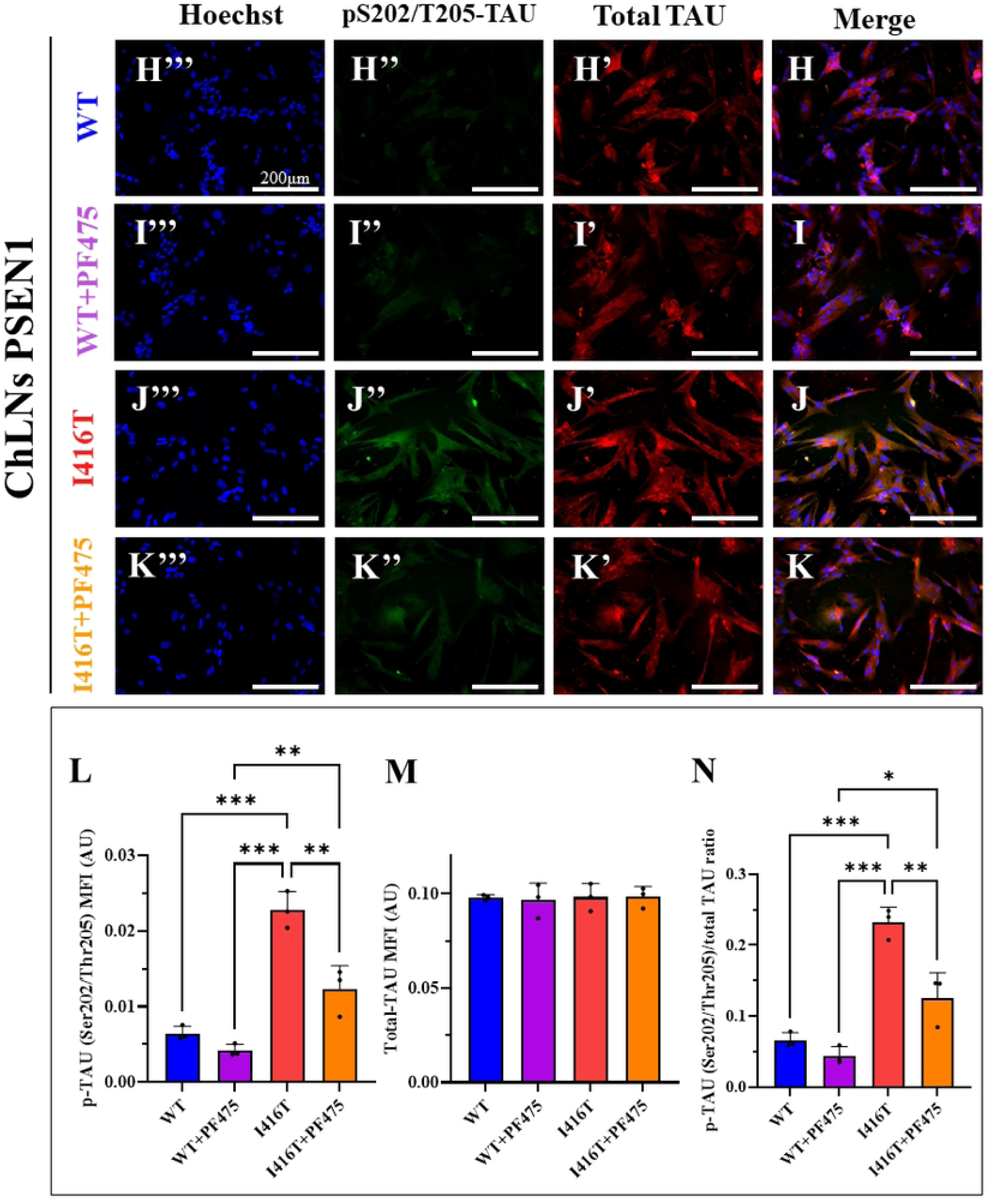
PF475 reduces pS202/T205-TAU protein levels in PSEN1 I416T Cholinergic-Like Neurons (ChLNs). WT PSEN1 and I416T MenSCs were cultured in a cholinergic differentiation medium for 7 days, followed by 4 days in regular medium in the absence or presence of PF475 (1 μM). (A-D) Representative flow cytometry density plot with density contours analysis showing phosphorylated TAU-positive cells at residue S202/T205 (A-D), total TAU-positive cells (A’-D’) in WT PSEN1 (A, B) or PSEN1 I416T ChLNs (C, D) in the absence (A, C) or presence (B, D) of PF475. (E-G) Quantification of pS202/T205-TAU-positive cells (E), total TAU-positive cells (F), and the pS202/T205/total TAU ratio (G). (H-K) Representative immunofluorescence merge image showing pS202/T205-TAU in WT (H, I) or PSEN1 I416T ChLNs (J, K) in the absence (H, J) or presence (I, K) of PF475 stained with antibodies against total TAU (H’-K’, red fluorescence), pS935-LRRK2 (H’’-K’’, green fluorescence). The nuclei were counterstained using Hoechst 33342 (H’’’-K’’’, blue fluorescence). (L-N) Mean fluorescence intensity (MFI) quantification of pS202/T205-TAU (L), total TAU (M), and the pS202/T205/total TAU ratio (N). Figures represent one of three independent experiments (n = 3). Results are represented as mean ± standard deviation. Statistical significance was analyzed using one-way ANOVA followed by Tukey’s test *p < 0.05, **p < 0.01, ***p < 0.001. Images were taken at 200× magnification.

### PF475 increases mitochondrial membrane potential (ΔΨ_m_) and reduces reactive oxygen species (ROS) generation in PSEN1 I416T ChLNs

Previous findings have shown that PSEN1 I416T ChLNs exhibit loss of ΔΨm and high production of ROS [11]. Therefore, we wanted to evaluate the effect of PF475 on mitochondria and on ROS. Indeed, PSEN1 I416T ChLNs decreased ΔΨm by −37% (Figure 5A) and produced ROS by +800% (Figure 5C) compared to WT ChLNs (Figures 5B and 5D). When exposed to PF475, it was harmless to WT ChLNs (Figures 5A-D), but induced an increase of ΔΨm by +45% (Figures 5A and 5B) and significantly reduced the generation of ROS by −83% in mutant neurons (Figures 5C and 5D). Similar observations were documented by fluorescence microscopy analysis (Figures 5E-J).

**Figure 5.**
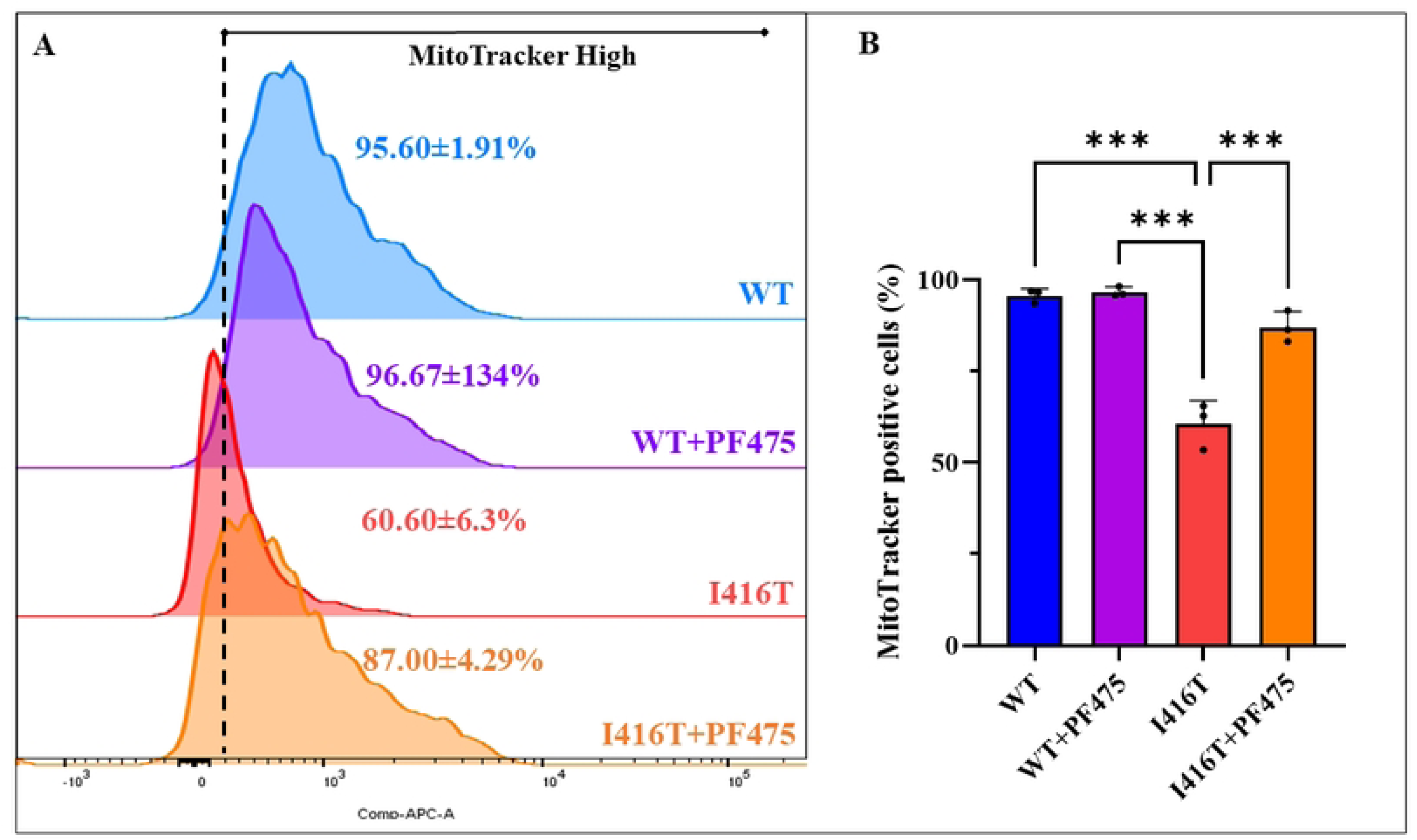
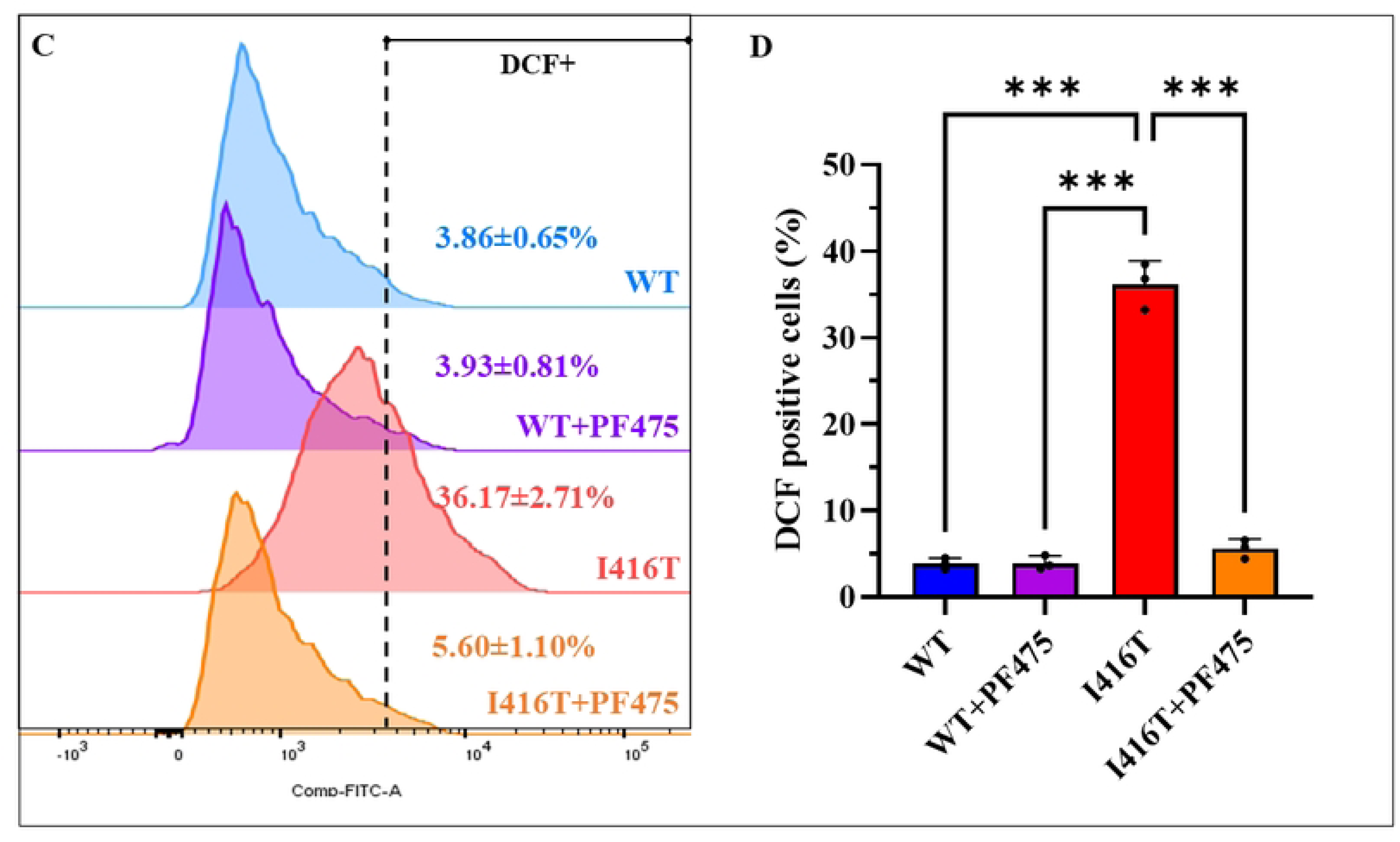
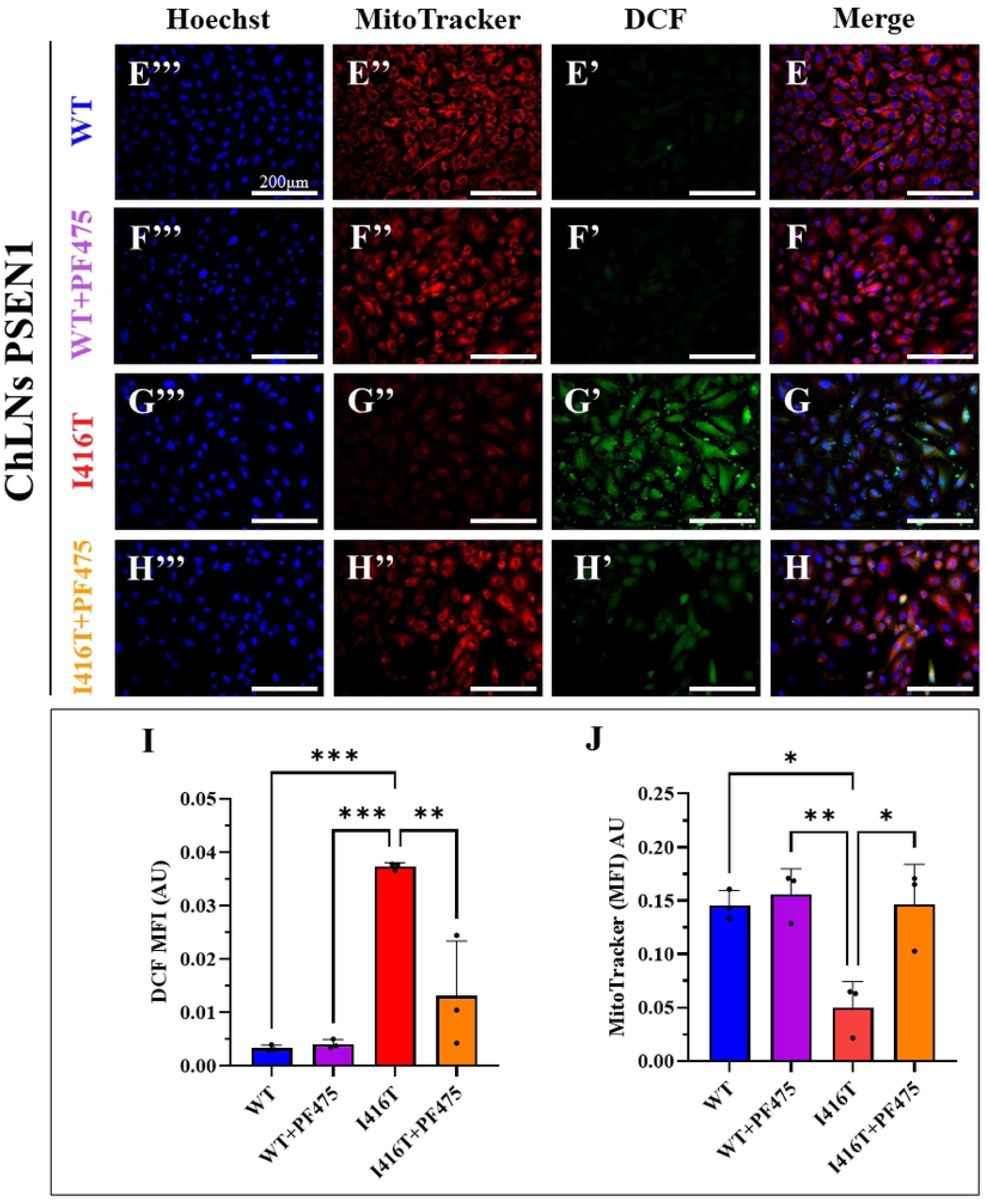
PF475 increases mitochondrial potential (ΔΨm) and blocks ROS accumulation in PSEN1 I416T Cholinergic-Like Neurons (ChLNs). WT PSEN1 and I416T MenSCs were cultured in a cholinergic differentiation medium for 7 days, followed by 4 days in regular medium in the absence or presence of PF475 (1 μM). (A) Representative flow cytometry histograms showing the percentage of cells with increased mitochondrial membrane potential (ΔΨm) in PSEN1 WT and I416T ChLNs in absence or presence of PF475. (B) Quantification of the percentage of MitoTracker-positive cells for the same groups. (C) Representative flow cytometry histograms showing ROS levels detected with DCF in PSEN1 WT and I416T ChLNs in absence or presence of PF475. (E-H) Representative immunofluorescence merge image showing DCF-positive and Mitotracker fluorescent cells in WT (E, F) or PSEN1 I416T ChLNs (G, H) in the absence (E, G) or presence (F, H) of PF475 stained with DCFH2-DA (E’-H’, green fluorescence), Mitotracker (E’’-H’’, red fluorescence). The nuclei were counterstained using Hoechst 33342 (E’’’-H’’’, blue fluorescence). (I-J) Mean fluorescence intensity (MFI) quantification of DFC (I), and Mitotracker (J). Figures represent one of three independent experiments (n = 3). Results are represented as mean ± standard deviation. Statistical significance was analyzed using one-way ANOVA followed by Tukey’s test *p < 0.05, **p < 0.01, ***p < 0.001. Images were taken at 200× magnification.

### PF475 diminishes cell death signaling transcription factor TP53 and PUMA protein in PSEN1 I416T ChLNs

Since PSEN1 I416T endogenously activates TP53 and PUMA [11], we investigated whether LRRK2 might be involved in the activation of these signaling molecules. To this end, WT and PSEN1 ChLNs were left untreated or treated with PF475. As shown in Figure 6, TP53 (Figure 6A) and PUMA (Figure 6F) were expressed at basal levels in untreated WT neurons and were not affected by the presence of PF475 (Figures 6B, 6E, 6G, and 6J). However, mutant ChLNs endogenously expressed TP53 (Figure 6C) and PUMA (Figure 6H) by +540% and +550%, respectively, in PSEN1 I416T compared to untreated WT ChLNs (Figures 6E and 6J). Upon treatment with PF475, TP53 (Figure 6D) and PUMA (Figure 6I) were reduced by −59% and −64%, respectively, in PSEN1 I416T ChLNs (Figures 6E and 6J). Similar results were obtained by fluorescence microscopy analysis (Figures 6K-P).

**Figure 6.**
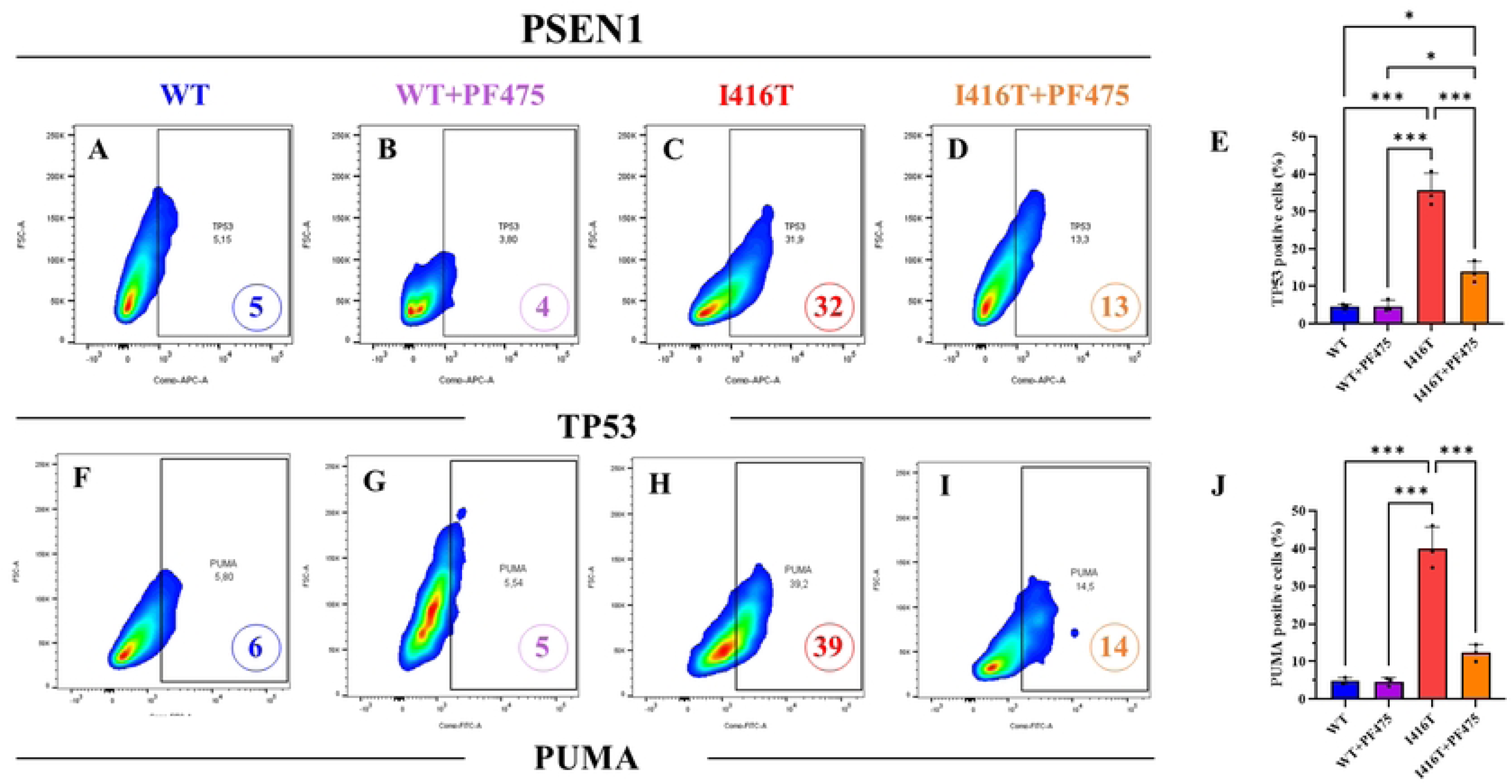
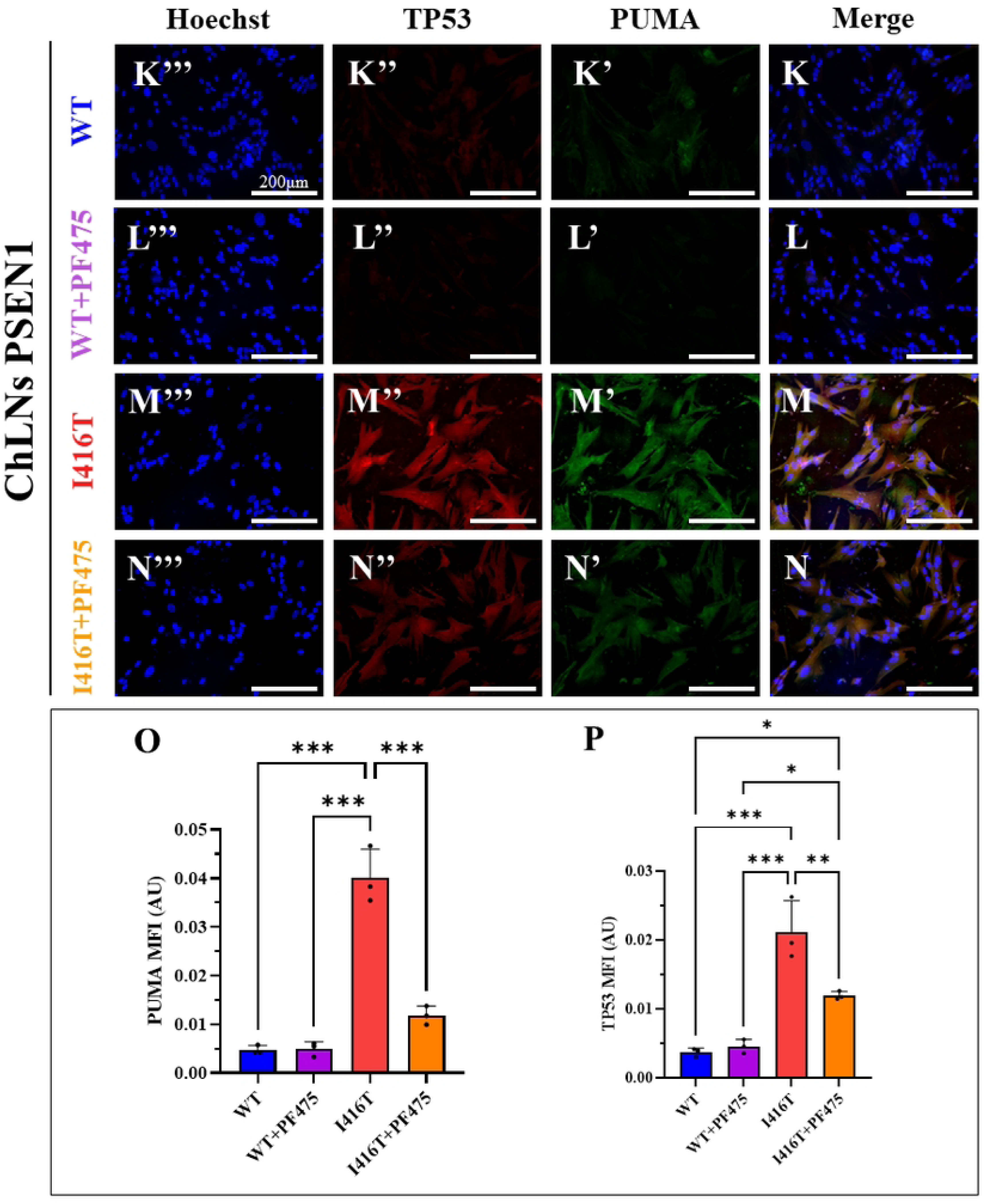
PF475 mitigates cell death signaling by reducing TP53 and PUMA expression in PSEN1 I416T Cholinergic-Like Neurons (ChLNs). WT PSEN1 and I416T MenSCs were cultured in a cholinergic differentiation medium for 7 days, followed by 4 days in regular medium in the absence or presence of PF475 (1 μM). (A-D) Representative flow cytometry density plot with density contours analysis showing TP53- positive cells in WT PSEN1 (A, B) or PSEN1 I416T ChLNs (C, D) in the absence (A, C) or presence (B, D) of PF475. (E) Quantification of TP53. (F-I) Representative flow cytometry density plot with density contours analysis showing PUMA-positive cells in WT PSEN1 (F, G) or PSEN1 I416T ChLNs (H, I) in the absence (F, H) or presence (G, I) of PF475. (J) Quantification of PUMA. (K-N) Representative immunofluorescence merge image showing PUMA and TP53 in WT (K, L) or PSEN1 I416T ChLNs (M, N) in the absence (K, M) or presence (L, N) of PF475 stained with antibodies against PUMA (K’-N’, green fluorescence), or TP53 (K’’-N’’, red fluorescence). The nuclei were counterstained using Hoechst 33342 (K’’’-N’’’, blue fluorescence). (O-P) Mean fluorescence intensity (MFI) quantification of PUMA (O) and TP53 (P). Figures represent one of three independent experiments (n = 3). Results are represented as mean ± standard deviation. Statistical significance was analyzed using one-way ANOVA followed by Tukey’s test **p < 0.01, ***p < 0.001. Images were taken at 200× magnification.

### PF475 decreases pro-death transcription factor c-JUN and executor protein cleaved caspase 3 (CC3)

In addition, we tested whether PF475 could also block activation of c-JUN and caspase 3, referred to as CC3 [11]. Figure 7 shows that untreated or treated WT expressed similar low basal levels of p-c-JUN at pathological Ser63/73 (Figures 7A and 7B) and inactive CC3 (Figures 7F and 7G), respectively. While PSEN1 I416T expressed p-c-JUN (Figure 7C) and CC3 (Figure 7H) by +617% and +620%, respectively, compared to untreated WT neurons (Figures 7E and 7J), the inhibitor decreased both markers by −58% (Figure 7D) and −83% (Figure 7I), respectively, compared to untreated mutant cells (Figures 7E and 7J). Fluorescence microscopy analysis showed similar results (Figures 7K-P).

**Figure 7.**
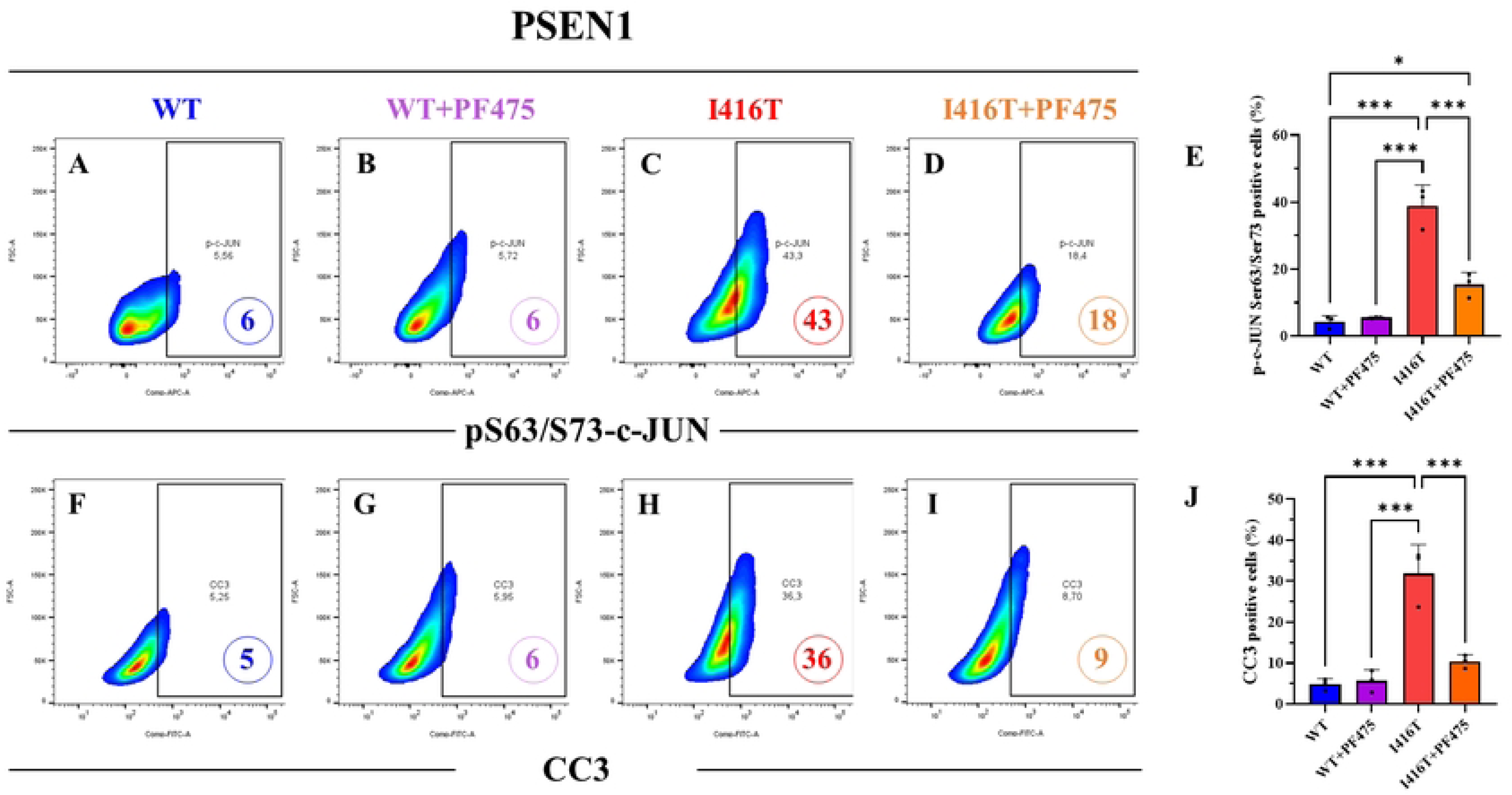
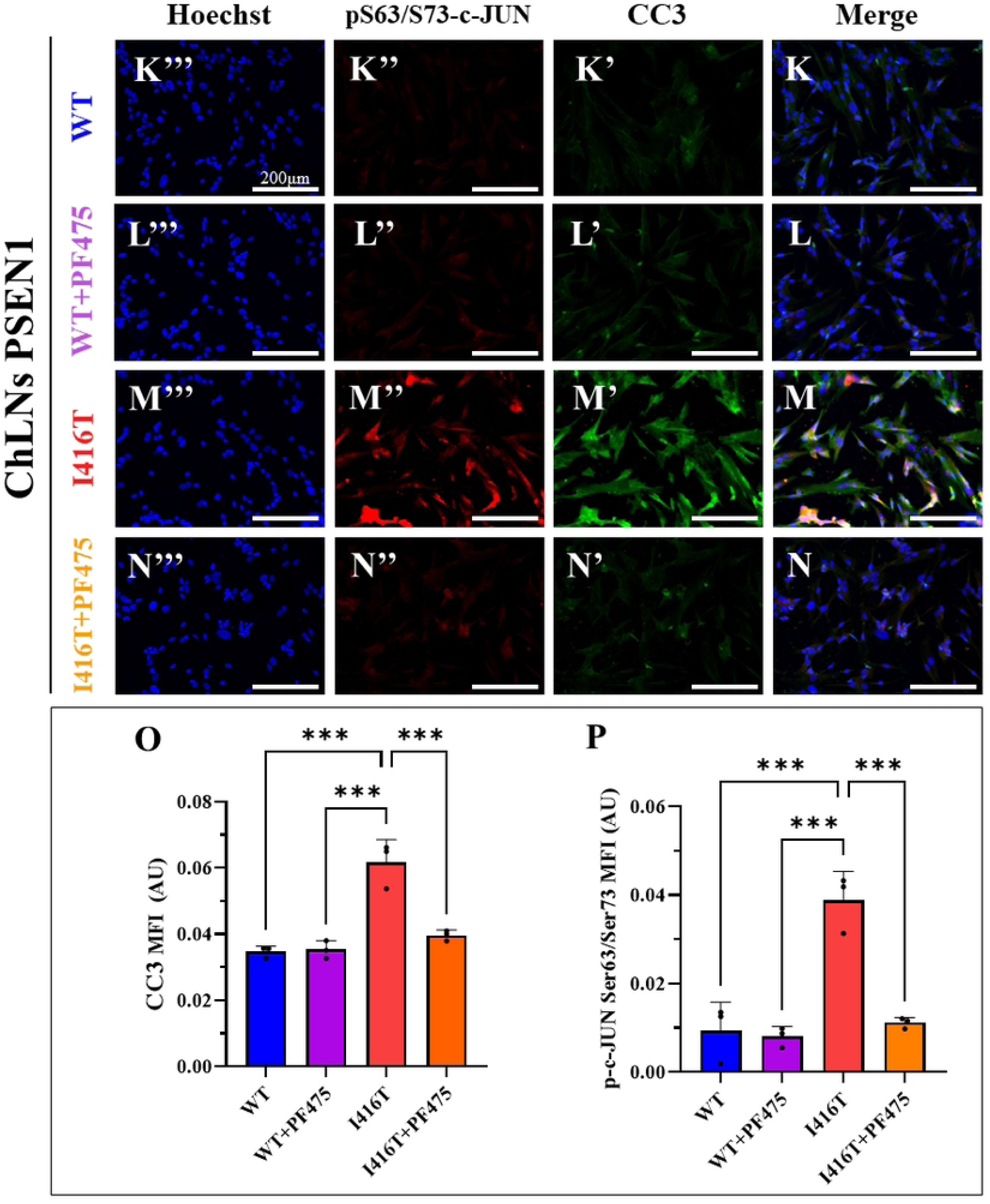
PF475 reduces pro-death transcription factor c-JUN and executor protein cleaved caspase 3 (CC3) in PSEN1 I416T Cholinergic-Like Neurons (ChLNs). WT PSEN1 and I416T MenSCs were cultured in a cholinergic differentiation medium for 7 days, followed by 4 days in regular medium in the absence or presence of PF475 (1 μM). (A-D) Representative flow cytometry density plot with density contours analysis showing pS63/73-c-JUN-positive cells in WT PSEN1 (A, B) or PSEN1 I416T ChLNs (C, D) in the absence (A, C) or presence (B, D) of PF475. (E) Quantification of pS63/73-c-JUN. (F-I) Representative flow cytometry density plot with density contours analysis showing CC3- positive cells in WT PSEN1 (F, G) or PSEN1 I416T ChLNs (H, I) in the absence (F, H) or presence (G, I) of PF475. (J) Quantification of CC3. (K-N) Representative immunofluorescence merge image showing CC3 and pS63/73-c-JUN in WT (K, L) or PSEN1 I416T ChLNs (M, N) in the absence (K, M) or presence (L, N) of PF475 stained with antibodies against CC3 (K’-N’, green fluorescence), or pS63/73-c-JUN (K’’-N’’, red fluorescence). The nuclei were counterstained using Hoechst 33342 (K’’’-N’’’, blue fluorescence). (O-P) Mean fluorescence intensity (MFI) quantification of CC3 (O) and pS63/73-c-JUN (P). Figures represent one of three independent experiments (n = 3). Results are represented as mean ± standard deviation. Statistical significance was analyzed using one-way ANOVA followed by Tukey’s test **p < 0.01, ***p < 0.001. Images were taken at 200× magnification.

### PF475 relieves PSEN1 I416T-induced dysfunctional autophagy lysosomal pathway (ALP) in ChLNs

Accumulating evidence suggests that KO or mutations in PSEN1 affect the lysosomal autophagy pathway [35, 36]. Therefore, we investigated the effect of PF475 on APL in PSEN1 I416T. For this purpose, WT and mutant ChLNs were left untreated or treated with PF475. We then evaluated lysosomal vacuole acidification with Lysotracker® reagent and autophagosome development with up/down levels of the autophagosome marker LC3-II (phosphatidylethanolamine-conjugated microtubule-associated protein 1A/1B-light chain 3 [37]). As shown in Figure 8, untreated WT ChLNs showed a basal level of autophagosome development as evidenced by 7% LC3-II marker (Figure 8A), whereas PSEN1 ChLNs showed an increase in LC3-II marker of +243% (Figure 8B), reflecting the accumulation of autophagosomes. Interestingly, WT ChLNs treated with PF475 increased LC3-II by +57% (Figure 8C), reflecting the enhancement of autophagosome development, but mutant ChLNs treated with PF475 dramatically reduced LC3-II by −45% (Figure 8D). For illustrative purposes, rapamycin (RAP), a positive inducer of autophagy [38], and bafilomycin A1 (BAF), a selective inhibitor of lysosomal vATPase [39], were included as control compounds. As expected, RAP increased LC3-II by +114% (Figure 8E), but BAF dramatically induced LC3-II accumulation by +628% in WT ChLNs (Figures 8F and 8G). Similar observations were made by fluorescence microscopy analysis (Figures 8H-N). Analysis of acidic vesicles (i.e., lysosomes) using Lysotracker® reagent shows that WT ChLNs had a basal percentage of Lysotracker-positive cells of 20% (Figure 9A) and that the mutant ChLNs reduced the percentage of Lysotracker-positive cells by −35% (Figure 9B) compared to untreated WT ChLNs. Figure 9C shows that PF475 had no effect on WT ChLNs (Figure 9C); however, mutant ChLNs increased lysotracker-positive cells to almost control levels when exposed to this inhibitor (Figure 9D). As expected, WT ChLNs exposed to RAP showed no significant change in the acidification of vacuoles (lysosomes, Figure 9E), but ChLNs exposed to BAF significantly decreased the detection of lysotracker-positive cells by −122% (Figures 9F and 9G). Fluorescence microscopy analysis revealed similar results (Figures 9H-N).

**Figure 8.**
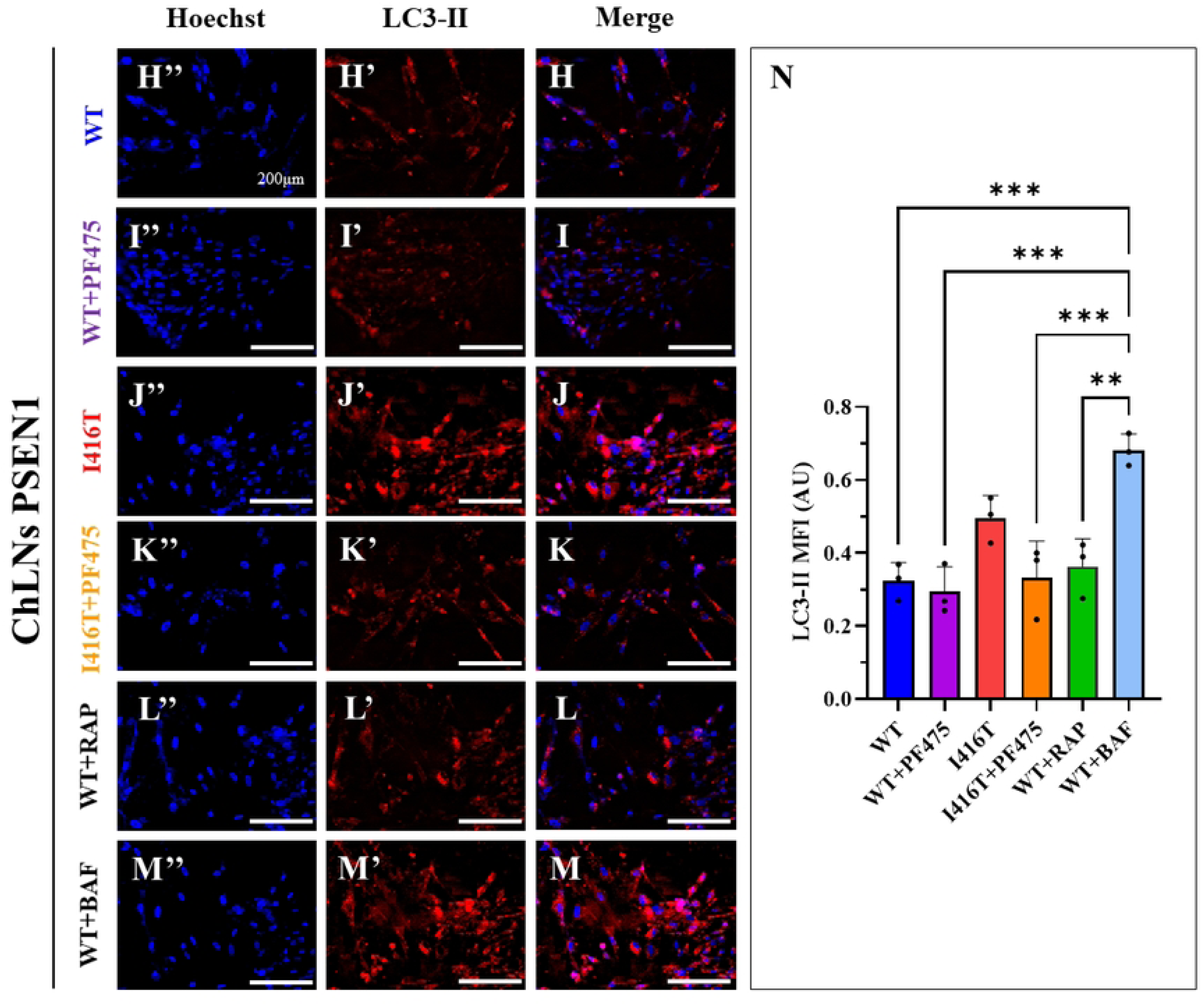
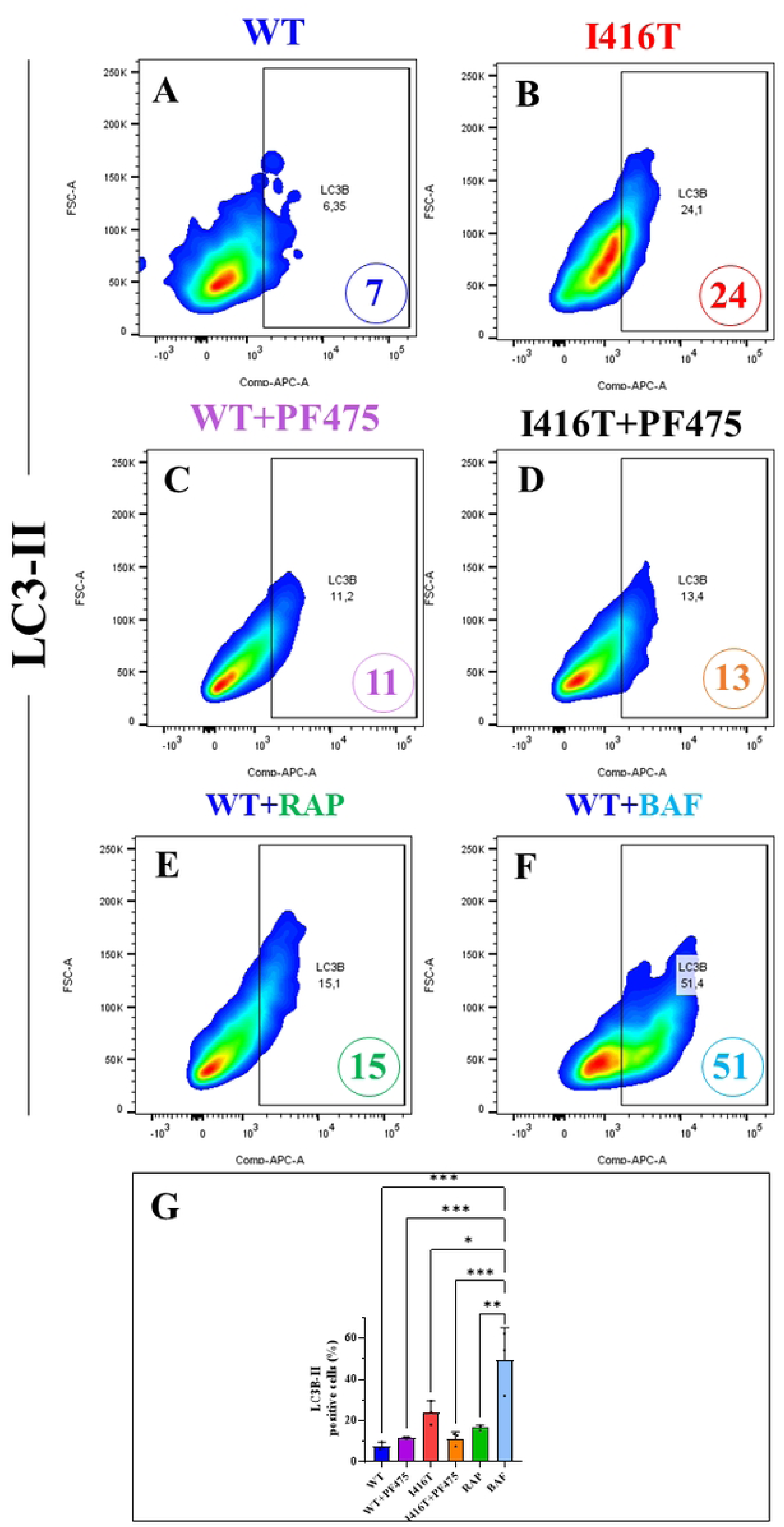
PF475 restores autophagy-lysosomal pathway functionality in PSEN1 I416T Cholinergic-Like Neurons (ChLNs). WT PSEN1 and I416T MenSCs were cultured in a cholinergic differentiation medium for 7 days, followed by 4 days in regular medium in the absence or presence of PF475 (1 μM). (A-D) Representative flow cytometry density plot with density contours analysis showing LC3- II-positive cells in WT PSEN1 (A, B) or PSEN1 I416T ChLNs (C, D) in the absence (A, C) or presence (B, D) of PF475, and WT cells treated with either rapamycin (1 nM RAP, E) or bafilomycin (1 nM BAF, F). Quantification of LC3-II (G). (H-M) Representative immunofluorescence merge image showing LC3-II in WT (H, I) or PSEN1 I416T ChLNs (J, K) in the absence (H, J) or presence (I, K) of PF475, and WT cells treated with either rapamycin (RAP, L) or bafilomycin (BAF, M) stained with antibodies against LC3-II (K’- N’, red fluorescence). The nuclei were counterstained using Hoechst 33342 (H’’-M’’, blue fluorescence). (N) Mean fluorescence intensity (MFI) quantification of LC3-II. Figures represent one of three independent experiments (n = 3). Results are represented as mean ± standard deviation. Statistical significance was analyzed using one-way ANOVA followed by Tukey’s test *p < 0.05, **p < 0.01, ***p < 0.001. Images were taken at 200× magnification.

**Figure 9.**
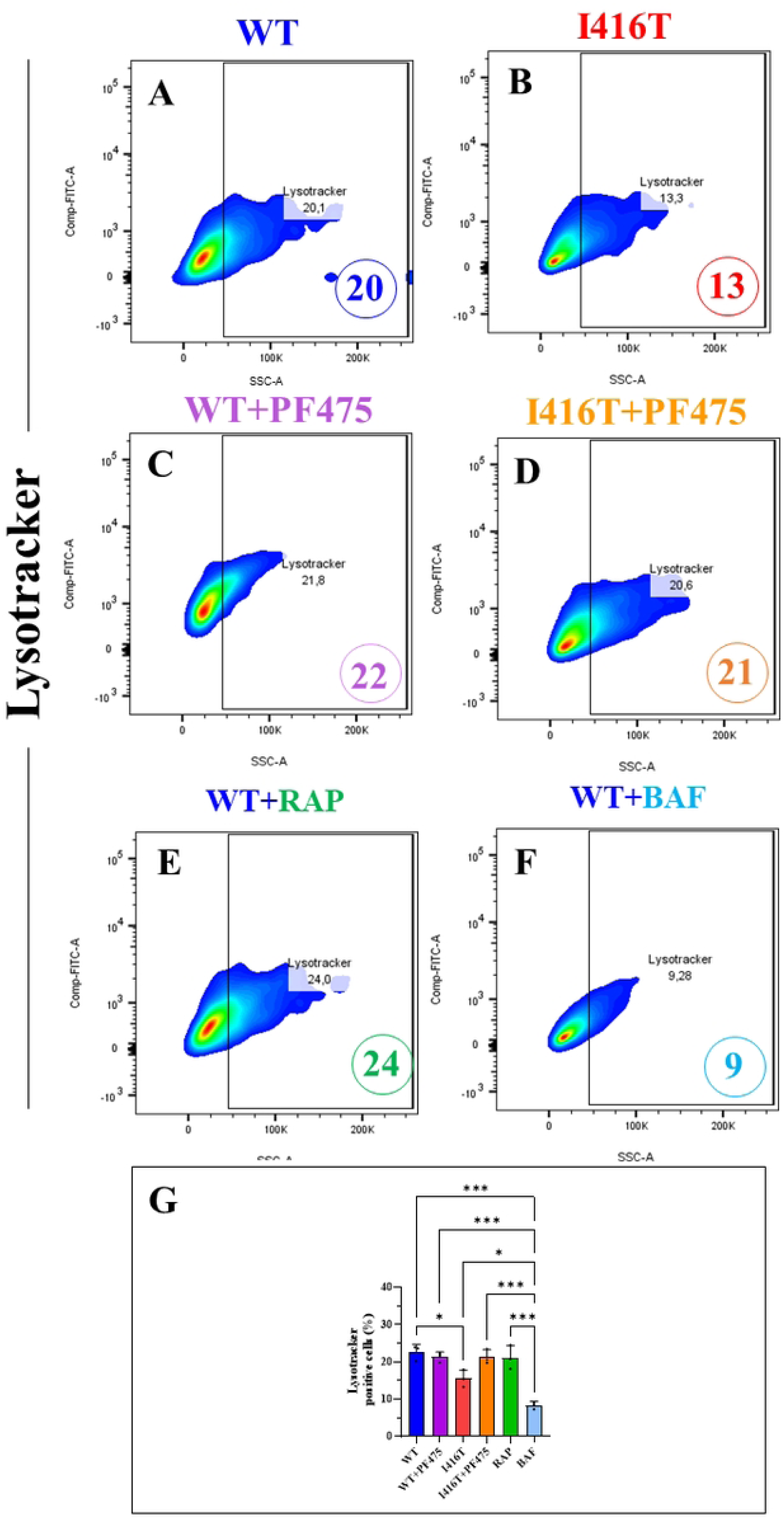
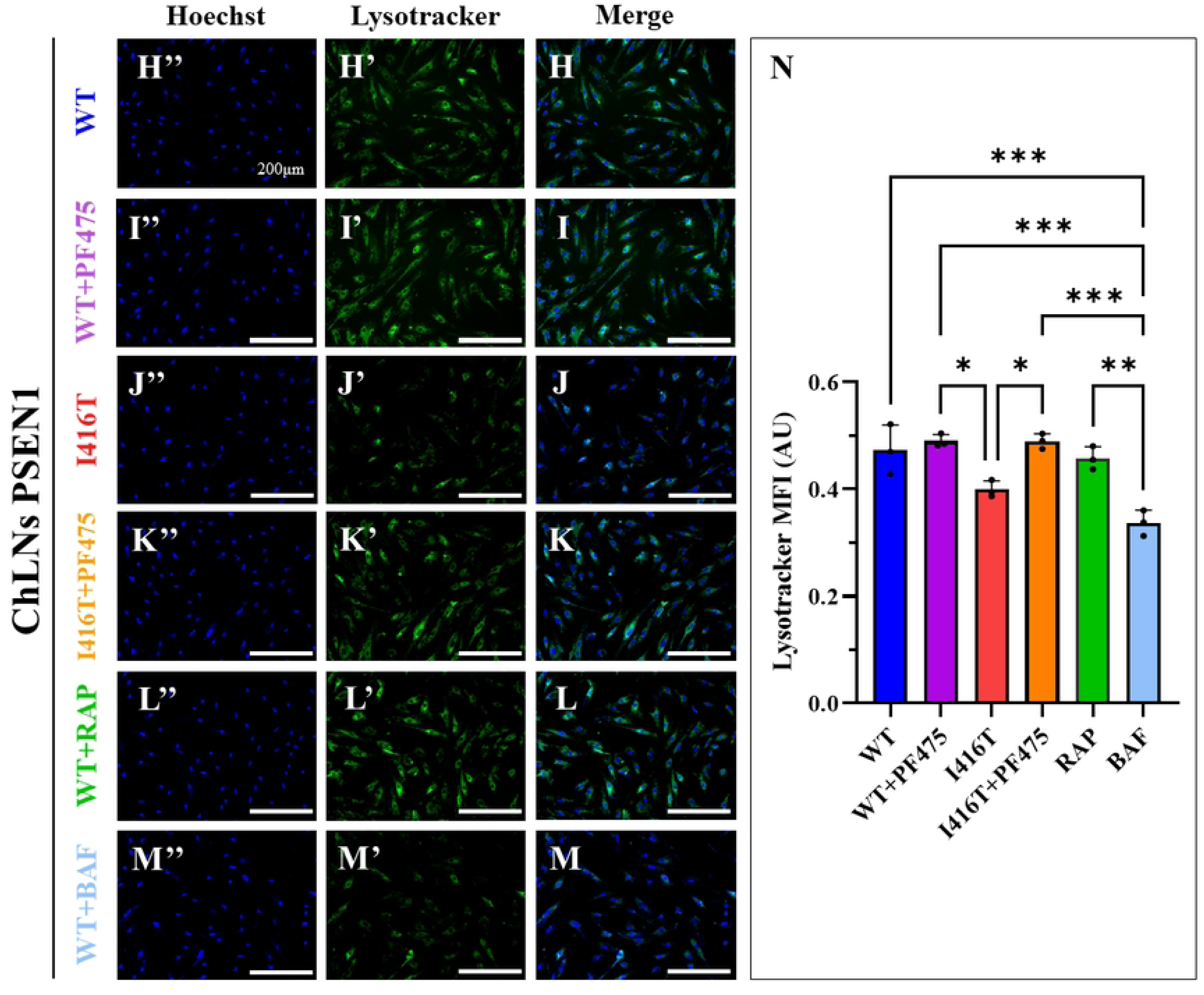
PF475 restores lysosomal pathway functionality in PSEN1 I416T Cholinergic-Like Neurons (ChLNs). WT PSEN1 and I416T MenSCs were cultured in a cholinergic differentiation medium for 7 days, followed by 4 days in regular medium in the absence or presence of PF475 (1 μM). (A-D) Representative flow cytometry density plot with density contours analysis showing Lysotracker-positive cells in WT PSEN1 (A, B) or PSEN1 I416T ChLNs (C, D) in the absence (A, C) or presence (B, D) of PF475, and WT cells treated with either rapamycin (1 nM RAP, E) or bafilomycin (1 nM BAF, F). Quantification of Lysotracker (G). (H-M) Representative fluorescence merge image showing Lysotracker in WT (H, I) or PSEN1 I416T ChLNs (J, K) in the absence (H, J) or presence (I, K) of PF475, and WT cells treated with either rapamycin (RAP, L) or bafilomycin (BAF, M) stained with Lysotracker (K’-N’, red fluorescence). The nuclei were counterstained using Hoechst 33342 (H’’-M’’, blue fluorescence). (N) Mean fluorescence intensity (MFI) quantification of Lysotracker. Figures represent one of three independent experiments (n = 3). Results are represented as mean ± standard deviation. Statistical significance was analyzed using one-way ANOVA followed by Tukey’s test *p < 0.05, **p < 0.01, ***p < 0.001. Images were taken at 200× magnification.

## Discussion

In the present investigation, we confirm that PSEN1 I416T ChLNs displayed the typical pathological phenotype of FAD, including accumulation of iAβ, generation of ROS, oxidation of the OS sensor protein DJ-1, pS202/S205-TAU, as well as loss of ΔΨm, and expression of the apoptosis markers TP53, p-S63/73-c-JUN, and CC3 [11]. Interestingly, similar to PSEN1 E280A [15], mutant I416T neurons also exhibited endogenous pS935-LRRK2 and pS129-αSyn. Our data suggest that LRRK2 may be a common kinase involved in the pathophysiology of AD and PD. However, whether pS935-LRRK2/pS129-αSyn contributes to the deterioration of cholinergic neurons in brain of FAD patients remains to be elucidated and merits further investigation. In addition, we report for the first time that mutant ChLNs exhibit abnormal ALP. In fact, I416T ChLNs are biologically similar to WT ChLNs treated with BAF. Indeed, we found that mutant ChLNs increased the accumulation of autophagosomes according to the LC3-II assay, but lysosome detection according to Lysotracker® was limited in I416T ChLNs. These observations suggest that the I416T mutation most likely affects the functionality of lysosomes by decreasing their lysosomal pH to a more basic rather than acidic vesicular environment through the loss of V0a1 vATPase subunits, which are normally involved in the transport of H^+^ to lysosomal vesicles [35]. Collectively, PSEN1 variants could function through PSEN/γ secretase-dependent or -independent pathways, as shown in Figure 10A. Importantly, iAβ induces apoptosis either through H₂O₂-induced signal transduction pathways [40] or through mitochondrial damage [41]. In fact, Aβ-induced H₂O₂ acts as a second messenger molecule [42], directly or indirectly involved in the oxidation of several proteins, including DJ-1 [43], activation of transcription factors (e.g, JNK/c-JUN) [44], and kinases such as LRRK2 (p-LRRK2, e.g., through the IKK pathway) [45], ultimately leading to neuronal cell death [46]. Accumulating evidence suggests that once activated, pS935-LRRK2 may function as a master regulator kinase involved in apoptosis and OS associated with PD [17, 18, 26, 47]. Indeed, in addition to directly or indirectly phosphorylating α-Syn [16, 48], p-LRRK2 has been shown to phosphorylate at least three key target molecules. (i) It activates MKK4, which activates the pro-apoptotic factor c-JUN via, for example, the MKK4/JNK pathway, MKK4/JNK pathway [49]; (ii) activates the mitochondrial fission dynamin-like protein 1 (DLP-1) protein, which together with the fission protein-1 (Fis-1) receptor [50] induces mitochondrial depolarization, fragmentation, and aggregation [51]; (iii) inactivates peroxiredoxin 3 (PRDX3) [52], thereby preventing H_2_O_2_ catalysis. We therefore postulate that abnormal LRRK2 kinase activity may also play a critical role in AD. Indeed, Henderson and coworkers [53] have demonstrated abundant Aβ pathology, prominent pS202/T205-TAU, and pS129-αSyn pathology in 12 brain samples from LRRK2 mutation carriers. Taken together, these observations suggest that p-LRRK2 and αSyn PD-associated pathology are consistent with comorbid AD pathology.

**Figure 10.**
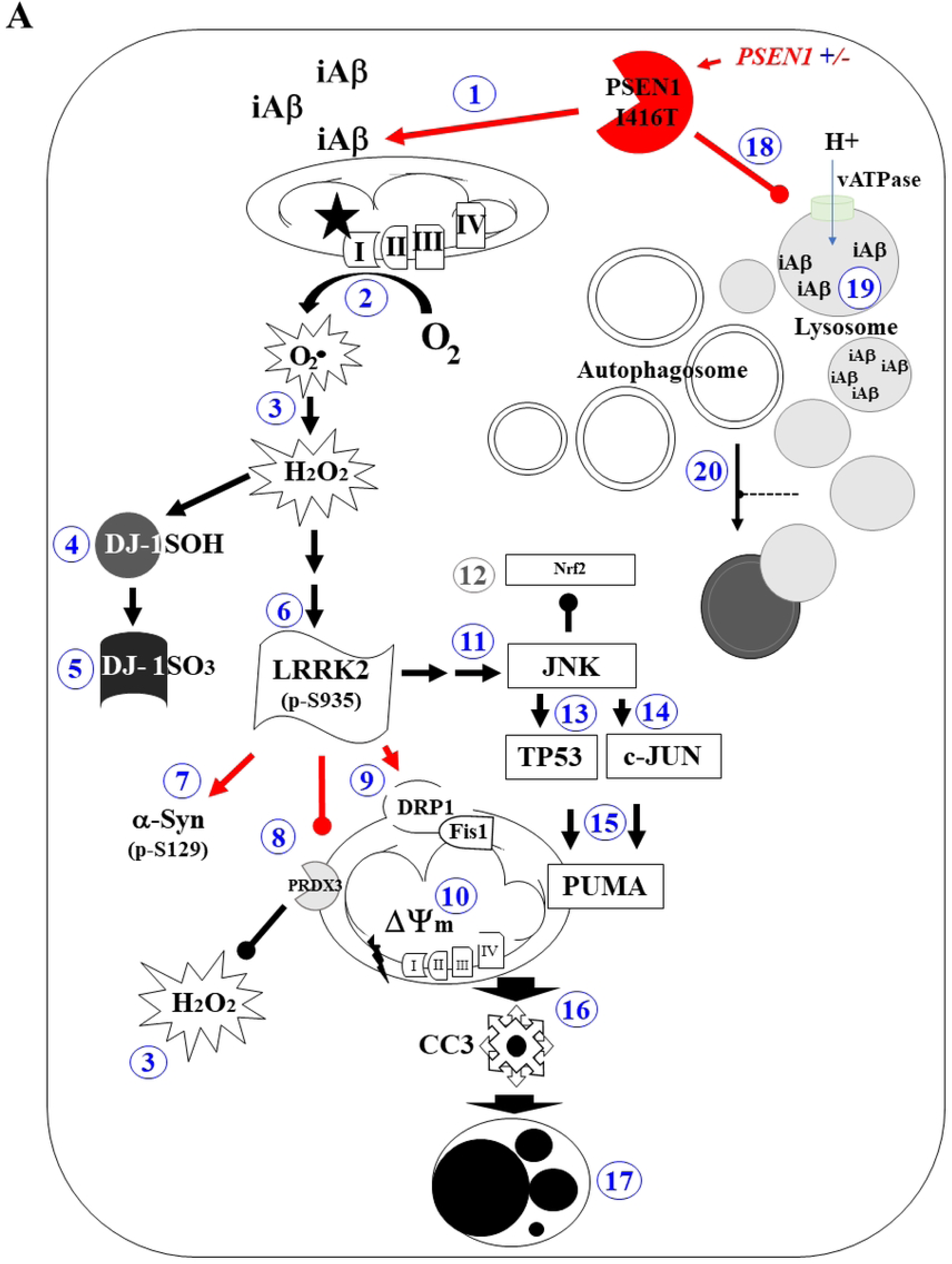
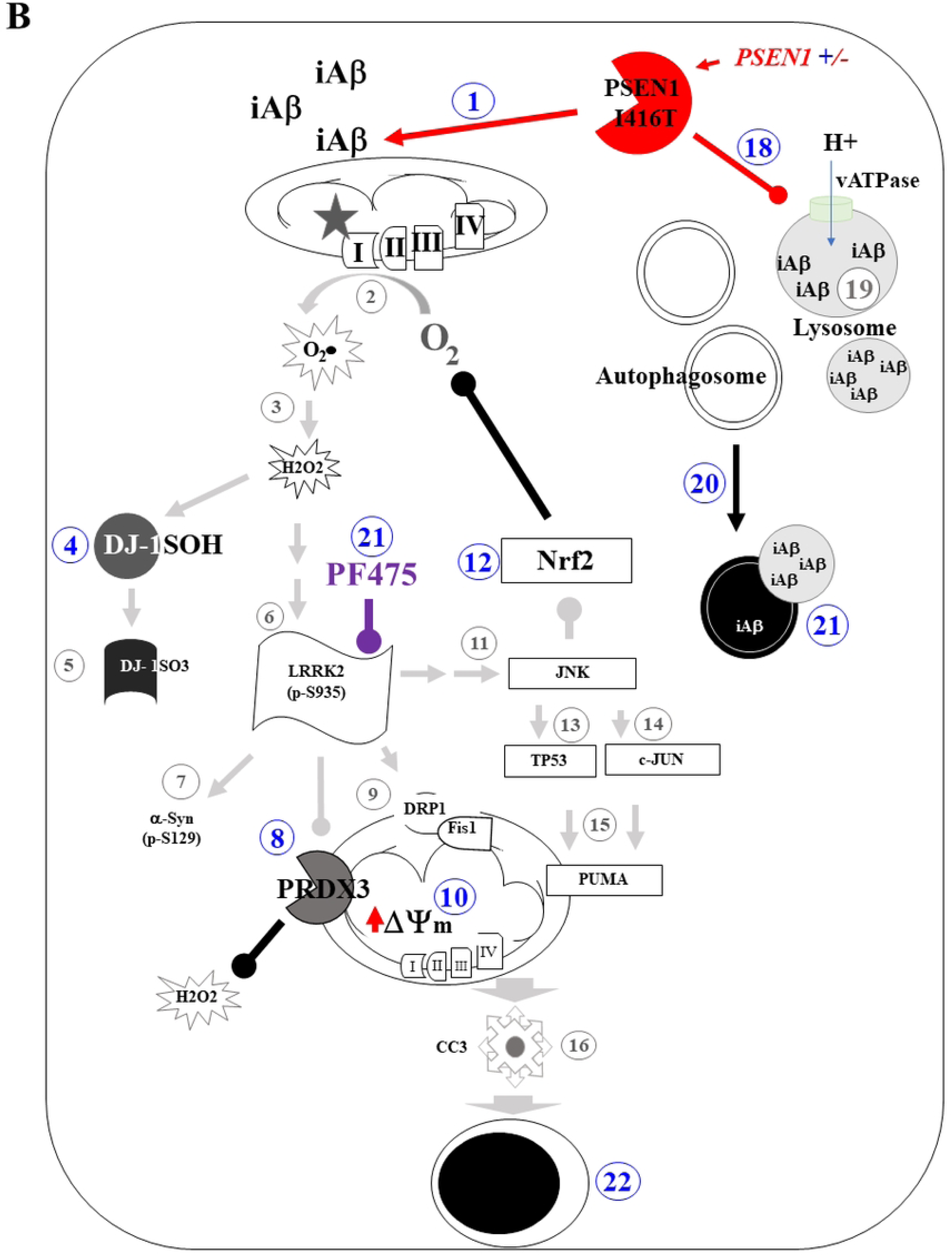
Schematic model of cell signaling induced by PSEN1/γ-secretase and effect of PF475 in PSEN1 I416T ChLNs. **(A)** *PSEN1/γ secretase-dependent signaling mechanism*. The PSEN1+/- gene encodes a 467 aa (catalytic and) non-catalytic PSEN1 I416T protein that, by a mechanism not yet fully understood, alters the metabolism of the type I transmembrane protein APP by overproducing iAβ (**1**). The iAβ peptide binds to the ubiquinone binding site of mitochondrial complex I (NADH: ubiquinone oxidoreductase), preventing electron transfer via flavin mononucleotide (FMN) to coenzyme Q10 (**2**). Interruption of the electron transport chain simultaneously generates an anion, superoxide (O₂^.-^), and hydrogen peroxide (H₂O₂, **3**). The latter is capable of oxidizing the stress sensor protein DJ-1Cys106-SH (**4**) to DJ-1Cys106-SO_3_ (**5**), which directly activates LRRK2 (leucine-rich repeat kinase 2) kinase through autophosphorylation (**6**). Once LRRK2 is phosphorylated at S935, the active p-S935 LRRK2 kinase phosphorylates four major targets: (i) (i) alpha-synuclein (αSyn) at the pathological residue S129 (**7**), (ii) inactivates protein peroxiredoxin 3 (PRDX3, **8**), preventing H_2_O_2_ catalysis; and (iii) activates the mitochondrial fission protein DLP-1 (dynamin-like protein 1, **9**), which, together with the fission protein-1 (Fis-1) receptor, induces loss of mitochondrial potential (ΔΨm, **10**). Finally, (iv) pS935-LRRK2 indirectly phosphorylated JNK (**11**), which in turn repressed Nrf2-associated expression of antioxidant proteins (**12**) and activated the transcription factors TP53 (**13**) and c-JUN (**14**). Both factors transcribe PUMA (**15**), a BH-3-only Bcl-2 protein involved in further mitochondrial depolarization (**10**). Impairment of mitochondrial potential leads to the release of apoptogenic proteins (e.g., cytochrome C), resulting in the production of cleaved caspase 3 (**16**), which is responsible for chromatin condensation and DNA fragmentation (**17**), typical apoptotic features in PSEN1 I416T ChLNs. **PSEN1/γ-secretase-independent signaling mechanism** Alternatively, PSEN1 binds directly to the V0a1 subunit of v-ATPase, affecting its maturation and assembly of the v-ATPase complex (**18**). The reduced catalytic activity of v-ATPase leads to an decrease in lysosomal pH, thereby limiting enzymatic degradation of obsolete components of the cell itself or toxic elements, such as Aβ (**19**). In addition, the fusion of autophagosomes and lysosomes is impaired (**20**), leading to their respective accumulation in mutant ChLNs. **(B)** Upon exposure to PF475 (**21**), p-LRRK2-associated apoptosis signaling is drastically reduced (*faint black line*). As a result, reactivated PRDX3 activity (**8**) and increased expression of Nfr2-induced antioxidant proteins (**12**) decrease Aβ-induced ROS/H₂O₂ (2, 3), thereby increasing ΔΨm (**10**) and increasing degradation of iAβ (**20**) through compensatory high autophagy flux (**21**). All these actions lead to global neuronal recovery and survival of PSEN1 I416T ChLNs (**22**).

Interestingly, genetic ablation or pharmacological inhibition of LRRK2 has been shown to be protective against DAergic neurodegeneration in both in vitro and in vivo models [19, 20, 24–26]. Therefore, silencing LRRK2 may reduce or eliminate kinase-associated deleterious signaling and provide neurons with an opportunity to survive. Here, we show that the LRRK2 inhibitor PF475 attenuated proteinopathy, OS, apoptosis, and dysfunctional ALP in PSEN1 I416T ChLNs (Figure 10B). This premise is supported by several observations. First, as expected, PF475 inhibited the phosphorylation of p-LRRK2 at S935 in I416T ChLNs to a comparable percentage of phosphorylation observed in inhibitor-treated WT ChLNs. As a result, pS129-αSyn in I416T ChLNs also decreased to a basal percentage of phosphorylation. These observations suggest a direct relationship between pS935-LRRK2 and pS129-αSyn and may serve as a distinctive pathological marker in FAD. However, p-LRRK2 and p-αSyn have not been systematically investigated as neuropathological markers in AD/FAD. Therefore, for pathological analysis, both markers should be included as routine tests in future AD/FAD brain autopsy cases or searched for in autopsy cases in the brain bank with documented PSEN1 variants for qualitative and/or quantitative analysis. Second, PF475 efficiently reduced the accumulation of iAβ and oxDJ-1. Although the exact mechanism by which this inhibitor blocks Aβ and prevents oxidation of DJ-1 remains to be elucidated, PF475 may reduce iAβ by improving ALP, thereby increasing the clearance of Aβ fragments (see further in the Discussion). Consequently, it might also reduce Aβ-induced generation of ROS/H₂O₂ via mitochondrial complex I, which is one of the major sources of ROS/H₂O₂ [54].

In fact, a significant reduction in ROS/H_2_O_2_ could subsequently stop further H_2_O_2_-induced associated signaling, including pS935-LRRK2 and mitochondrial damage [46]. This observation may also explain why PF475 rescued ΔΨm in PSEN1 I416T ChLNs. Interestingly, pS935-LRRK2 indirectly activates JNK [49], which in turn phosphorylates and downregulates either nuclear factor erythroid 2-related factor 2 (Nfr2) [55], which is involved in xenobiotic detoxification and expression of cytoprotective proteins [56, 57], or inhibits PRDX3 activity [52]. Therefore, blocking p-LRRK2 might increase the expression of antioxidant proteins and enhance peroxireductase activity. Third, PF475 decreased pS202/T205-TAU. A possible explanation for this observation is that PF475 might break the Aβ driven JNK>p-TAU axis [58] by blocking LRRK2 [59]. Finally, as noted above, PF475 not only increased ΔΨm and reduced ROS, but it also significantly reduced the apoptotic executioner protein CC3 in I416T ChLNs. This result is not surprising given that the upstream deleterious signaling has already been eliminated. As a proof of concept, PF475 dramatically reduced pS63/S73-c-JUN, TP53 and PUMA. Thus, specific inhibition of p-LRRK2 by PF475 results in deactivation of the cell death cascade JNK> TP53, c-JUN> PUMA> loss of ΔΨm> CC3.

Several data suggest that PSEN1 is required for proper autophagy in neurons [60, 61]. Indeed, it was found that KO or mutations in PSEN1 altered substrate proteolysis and autophagosome clearance during autophagy by failing PS1-dependent targeting of the v-ATPase V0a1 subunit to lysosomes, resulting in selective impairment of autolysosome acidification [35]. Alternatively, PSEN1 deficiency may cause autophagy suppression in human neuronal stem cells through downregulation of the ERK/CREB pathway [62]. Whatever the mechanism, PSEN1 is essential for v-ATPase targeting to lysosomes and proteolysis during autophagy. Consistent with this view, PSEN1 I416T induced a significant accumulation of autophagosomes (i.e., as evidenced by a high percentage of LC3-II-positive cells) and decreased lysosomal acidification (as indicated by a low fluorescence signal according to the Lysotracker® assay), resulting in defective Aβ-dependent autophagy-lysosomal proteolysis, as reflected by the accumulation of iAβ in ChLNs. Notably, I416T-induced ALP defects mimic WT ChLNs upon BAF exposure. This observation suggests that the I416T mutation may be involved in the impairment of lysosomal acidification and Aβ proteolysis. Remarkably, PF475 restored the flow of autophagic phases by increasing the development of autophagosomes and lysosomal acidification. One possible explanation is that p-LRRK2 could decrease autophagosome-lysosome fusion and maintain lysosomal pH by directly binding to H^+^-ATPase [63]. However, LRRK2 is also involved in ALP, including regulation of phagophore and autophagosome formation, autophagosome/lysosome fusion, lysosomal maturation, calcium levels, and lysosomal protein degradation [21]. Whatever the mechanism, LRRK2 mutations may disrupt lysosomal function in a kinase-dependent manner. Therefore, specific inhibition of p-LRRK2 could restore autophagy-lysosomal activity in PSEN1 I416T ChLNs. Taken together, these findings suggest that PF475 increases autophagy flux and thereby detoxifies ChLNs from iAβ accumulation.

Despite several therapeutic attempts to treat AD, including immunotherapy [64] and non-drug therapies [65], they have either failed or shown limited efficacy. Part of the reason for these puzzling results may be the limited knowledge of Aβ pathophysiology (e.g., ref. [66]). Several data suggest that iAβ, rather than eAβ, may be the earliest insult involved in the destruction of cholinergic and/or hippocampal neurons in human AD brain [67–71], rodents [72, 73], and in vitro models of FAD [11, 27, 74]. Therefore, drug discovery efforts targeting iAβ or iAβ-induced signaling molecules should be the primary focus for future therapeutic strategies [46]. Here, we show that PF475 directly blocks p-LRRK2 and indirectly promotes the deactivation of pro-death proteins and normalization of ALP in mutant ChLNs. Thus, PF475 reveals LRRK2 as a potential druggable kinase in AD, as has been proposed for the treatment of PD [75]. Interestingly, radiolabeled [11C]PF06447475 has shown binding specificity for LRRK2, exhibiting high bound radioactivity in several rat brain regions, including the striatum, cerebral cortex, and hippocampus, whereas low bound radioactivity was detected in the pons and cerebellum [76]. Notably, these are the same regions implicated in AD [5]. However, whether PF475 could be useful for the treatment of FAD requires further validation in the clinic.

## Conclusion

Despite the cellular and molecular complexity of AD [77], we have identified for the first time LRRK2 as a master regulatory protein involved in cell death and abnormal ALP in PSEN1 I416T ChLNs. Indeed, LRRK2 kinase activity directly or indirectly modulates the accumulation of intracellular neurotoxic proteins (e.g., Aβ, pS202/S205-TAU, pS129-αSyn), generation of OS (e.g., ROS/H_2_O_2_; oxDJ-1), loss of ΔΨm, abnormal activation of apoptotic transcription factors (e.g., TP53, c-JUN), apoptogenic proteins (e.g., PUMA, CC3), accumulation of autophagosomes and alteration of acidic lysosomal vacuoles in PSEN1 I416T ChLNs (Figure 10A). Interestingly, PF475 significantly reduced proteinopathy and OS, restored ALP, and reduced apoptosis, thereby restoring the survival of mutant ChLNs (Figure 10B). Since LRRK2 kinase inhibitors are promising therapeutic drugs for the treatment of PD [78,79], our results suggest that LRRK2 may also be a potential target for therapeutic intervention in FAD.

Using MenSCs as a cellular FAD system model offers several advantages over other biological sources, such as hiPSC-derived neuronal models. Firstly, MenSCs can be obtained via a minimally invasive and safe procedure using a menstrual cup [80]. However, the menstrual sample must comply with level-2 biohazard requirements [81]. Importantly, this procedure is age-dependent for both volunteers and FAD patients. Secondly, MenSCs raise minimal bioethical concerns according to ethical review boards. Thirdly, MenSCs have been shown to be cellularly and biochemically equivalent to hiPSCs [27]. Fourthly, the methodology and MenSCs differentiation protocols are straightforward and cost-effective, saving laboratory time. Consequently, MenSCs-derived ChLNs can be obtained within seven days [30] and can reproduce PSEN1 I416T FAD pathology within 11 days [11]. In contrast, iPSC-derived ChNs are time-consuming and costly. The isolation and purification procedures (e.g., skin fibroblasts) are technically challenging, expensive, time-consuming, and labor-intensive. Furthermore, the protocol for differentiating iPSCs into neuronal progenitor cells (NPCs), which can then be guided to form different neuronal lineages, such as cholinergic neurons, takes at least 35 days [82]. Fifthly, the MenSC-derived ChLNs model mimics many features of brain cholinergic neurons, highlighting its utility in studying cholinergic function. Last but not least, MenSC-derived ChLNs provide an excellent platform for testing potential drug targets and candidates (e.g., in this study) in FAD. However, given the origin of MenSCs, the present observations and conclusions are limited to female gender. Accordingly, studies including male samples (e.g., Wharton’s jelly mesenchymal stem cells or dental pulp stem cells) are needed to confirm the universality of our observations.

## Author Contributions

Conceptualization: M.J.-D.-R. and C.V.-P.; methodology, M.J.-D.-R. and C.V.-P.; validation, N.G.-S.; formal analysis, N.G.-S. and C.V.P.; investigation, N.G.-S.; resources, M.J.-D.-R. and C.V.-P.; data curation, N.G.-S.; writing—original draft preparation, M.J.-D.-R. and C.V.-P.; writing—review and editing, N.G.-S., C.V.-P. and M.J.-D.-R.; visualization, M.J.-D.-R. and C.V-P.; supervision, M.J.-D.-R.; project administration, M.J.-D.-R.; funding acquisition, M.J.-D.-R. and C.V.-P. All authors have read and agreed to the published version of the manuscript.

## Institutional Review Board Statement

Menstrual specimen donors provided a signed informed consent and experimental protocols were approved by the ethics committee of the Sede de Investigación Universitaria (SIU), University of Antioquia, Medellín, Colombia (Act 2020-10854). All methods were carried out in accordance with relevant guidelines and research from Colombian regulations (Law 2253 of 2022 Congress of the Republic of Colombia).

## Informed Consent Statement

Informed consent was obtained from all subjects involved in the study.

## Clinical trial number

not applicable.

## Data Availability Statement

All data generated or analyzed during this study are included in this published article.

